# Maternal gut and vaginal microbiota, alcohol use during pregnancy, and infant fetal alcohol spectrum disorder diagnosis in a South African cohort

**DOI:** 10.64898/2026.08.04.26359606

**Authors:** Lauren C. Martin, Natasha Kitchin, Jacqueline S. Womersley, Kristien Nel Van Zyl, Anna-Susan Marais, Marlene M. de Vries, Matthew J. Dalby, Raymond Kiu, Lindsay J. Hall, Philip A. May, Soraya Seedat, Sian M.J. Hemmings

## Abstract

**Background:** The detrimental impact of alcohol consumption on the gut microbiome is well-established. However, less is known about how alcohol exposure during pregnancy affects the maternal gut and vaginal microbiota, or how these microbial changes relate to subsequent infant diagnosis of fetal alcohol spectrum disorder (FASD). We therefore investigated associations between self-reported alcohol use during pregnancy, infant FASD diagnosis, and maternal gut and vaginal microbiota.

**Methods:** Fecal samples (n = 207) and vaginal swabs (n = 28) from pregnant participants recruited through antenatal clinics in the Western Cape Province of South Africa were profiled by 16S rRNA V1-V2 amplicon sequencing. Maternal alcohol use was assessed using Alcohol Use Disorder Identification Test (AUDIT) scores, and FASD was diagnosed in infants using revised Institute of Medicine criteria. Microbial diversity, taxonomic profiles and *PICRUSt2*-predicted functional pathways were analyzed using *vegan*, *phyloseq* and *MaAsLin3*.

**Results:** Maternal AUDIT scores were negatively associated with maternal gut microbiota richness, Shannon, and Inverse Simpson diversity (*p* < 0.05). Observed richness was also reduced in participants whose infants were diagnosed with FASD (*p* = 0.046). Gut microbiota community structure was not significantly associated with alcohol use or infant FASD diagnosis. However, taxonomic and functional analysis suggested gram-positive taxa depletion with alcohol use and FASD diagnosis, and impaired one-carbon metabolism among participants with infants diagnosed with FASD. Vaginal microbiota diversity and composition were not associated with alcohol use or infant FASD diagnostic group.

**Conclusions:** This is the first human study to investigate the maternal gut and vaginal microbiota in relation to alcohol use during pregnancy and infant FASD outcomes. Our findings suggest that alcohol use is associated with disruption of the maternal gut microbiota, with potential implications for FASD development in exposed infants. Further investigation of alcohol-associated maternal microbial disturbances may inform microbiota-targeted strategies to improve maternal and infant health linked to alcohol use.

## Introduction

Prenatal alcohol exposure (PAE) is an established risk factor for adverse pregnancy outcomes (DeJong et al., 2019), and can alter the development, function, and regulation of fetal neural and physiological systems, manifesting its effects through a range of life-long physical abnormalities, and cognitive and behavioural deficits (Hellemans et al., 2010; Kane et al., 2012) falling under the umbrella term, fetal alcohol spectrum disorders (FASD) (Popova et al., 2023). Despite efforts to communicate the dangers of PAE, alcohol use during pregnancy remains a significant problem in South Africa, particularly among rural and disadvantaged communities (May et al., 2017), where the prevalence of FASD, reported to be 0.77% globally (Roozen et al., 2016), is estimated to be between 20.6 - 36.6% in the Western Cape Province (May et al., 2022).

Alcohol significantly impacts the diversity and structure of the gut microbiota. Reductions in the abundance of the typically dominant bacterial phyla *Bacillota* (formerly *Firmicutes*) and *Bacteroidota* (formerly *Bacteroidetes*), and enrichment of *Proteobacteria (*now *Pseudomonadota)* species have been documented in the context of alcohol use (Engen et al., 2015; Litwinowicz et al., 2020; Smith, 2025). Depletion of several bacterial genera, including *Akkermansia*, *Bacteroides*, *Clostridium*, *Collinsella*, *Faecalibacterium*, *Parabacteroides*, *Prevotella,* and *Ruminococcus* has also been reported, many of which contribute to short chain fatty acid (SCFA), branch-chained amino acid, bile acid, and tryptophan derivative production, and support intestinal barrier integrity and enteric immunity (Litwinowicz et al., 2020; Smith, 2025; Sosnowski and Przybyłkowski, 2024). Beyond alterations in composition, alcohol disrupts intestinal barrier function and promotes gut hyperpermeability, facilitating translocation of pro-inflammatory intestinal microbiota and metabolic products into circulation and contributing to systemic inflammation (Engen et al., 2015; Smith, 2025). Emerging evidence also suggests that alcohol may also destabilize “optimal”, health-promoting vaginal microbial communities dominated by *Lactobacillus crispatus*, *Lactobacillus gasseri*, and *Lactobacillus jensenii* species in favor of sub-optimal community states (Djusse et al., 2025; Kamiya et al., 2025; Morsli et al., 2024). These include *Lactobacillus iners-* dominated communities, or *Lactobacilli*-depleted states characterized by an overgrowth of facultative anaerobes, which confer greater susceptibility to sexually transmitted infections and poorer reproductive outcomes (Djusse et al., 2025; Kamiya et al., 2025; Morsli et al., 2024).

Though the immune, metabolic and hormonal alterations that support fetal development during pregnancy are known to shift microbiota composition and reduce bacterial richness across body sites (Nuriel-Ohayon et al., 2016), the impact of alcohol use during pregnancy remains comparatively underexplored, particularly with respect to the vaginal microbiota, for which no animal or human studies have been reported to date. Gut microbial diversity reductions, extending beyond pregnancy-associated losses, and depletion of *Lactobacillus*, *Oscillospiraceae* (formerly *Ruminococcaceae*), and *Lachnospiraceae* have been reported in alcohol-consuming dams (Bodnar et al., 2024), alongside altered community composition in their exposed offspring (Bodnar et al., 2024, 2022). Positive associations between maternal drinking frequency and *Phascolarctobacterium* and *Blautia,* and negative associations with *Faecalibacterium* have also been reported in humans (Wang et al., 2021). Collectively, these findings suggest that alcohol perturbs the maternal gut microbiota, and impacts early life offspring gut microbiome establishment (Bodnar et al., 2022, 2024; Wang and Lin, 2021).

Maternal gut microbiota are proposed to influence fetal development through the maternal-fetal circulation of microbiota and microbial metabolites, and maternal immune system modulation (Suárez-Martínez et al., 2023). Direct contact with and ingestion of maternal microbiota during delivery also contributes to neonatal gut microbiome establishment (Suárez-Martínez et al., 2023), which plays a fundamental role in early brain development (Sajdel-Sulkowska, 2023; Warner, 2019). As such, dysbiotic shifts in maternal microbiota composition may promote non-optimal neonatal gut microbiome colonization, contributing to impaired metabolic and digestive functions, and a heightened risk of behavioural dysregulation (Hillemacher et al., 2018), and neurodevelopmental (Warner, 2019) and mood disorders (Valles-Colomer et al., 2019). Alcohol-induced maternal gut and vaginal microbiota alterations may therefore increase the risk of FASD development.

No human studies have examined the maternal gut and vaginal microbiota in relation to alcohol consumption during pregnancy and infant FASD diagnosis. Here, we conducted a cross-sectional study examining the gut (n = 207) and vaginal (n = 28) microbiota of pregnant South African women to determine whether alcohol use during pregnancy alters maternal microbiome characteristics, and whether these features are associated with later alcohol-related neurodevelopmental disorders in their infants’. These findings expand our understanding of the detrimental impact of alcohol use during pregnancy and may inform microbiota-targeted intervention strategies with potential to improve maternal and infant health linked to alcohol use.

## Methods

### Study description

This study aimed investigates the gut and vaginal microbiota of pregnant South African women in relation to alcohol use during pregnancy and, in a subset of participants, infant FASD diagnosis. Our primary aim was to characterize gut (n = 207) and vaginal (n = 28) microbial community composition, diversity, and predicted functional potential in relation to self-reported alcohol use during pregnancy, examined as a continuous measure and a binary indicator of hazardous alcohol use (HAU). As a secondary exploratory aim, we examined these microbial features in a subset of participants whose infants underwent diagnostic assessment for FASD [gut (n = 168) and vaginal (n = 15)]. In line with current literature (Kamiya et al., 2025; Bodnar et al., 2024), we anticipated that alcohol exposure during pregnancy would be associated with reduced gut and greater vaginal microbial diversity.

### Study population

This study is an auxiliary pilot project to a National Institutes of Health (NIH) National Institute on Alcohol Abuse (NIAAA)-funded FASD epidemiological and clinical research study [R01/U01AA15134] that has integrated complementary multidisciplinary expertise to facilitate early-stage research in high-risk regions of the Western Cape of South Africa. This study was approved by the Stellenbosch University Health Research Ethics Committee (N13/01/013 and S19/07125). Written informed consent was obtained from all participants who were interviewed, assessed, and provided biological samples.

Pregnant women were recruited by the Fetal Alcohol Syndrome Epidemiology Research (FASER) team from primary health care clinics in the communities of Wellington and Robertson in the Western Cape province of South Africa. The study population originates from rural deciduous fruit and wine-producing communities of the Western Cape with the highest documented regional rates of FASD globally (May et al., 2022, 2017). Children in this population may be exposed to large amounts of alcohol prenatally, in frequent and regularly timed binges, which is rare in other studies of PAE (May et al., 2022).

Participants more than 38 weeks pregnant or with major pregnancy-related and/or other serious medical conditions, a history of active or uncontrolled gastrointestinal disorders *e.g.,* recent diarrhea, irritable bowel syndrome, inflammatory bowel disease or Crohn’s disease, or reported past-month antibiotic use were excluded. Any antibiotic use within the previous six months was noted.

### Maternal assessments

Clinical interviews, conducted during antenatal and subsequent postnatal visits, were undertaken by FASER team registered nurses or social workers and collected detailed information including demographic, general health, pregnancy details, childbearing history, alcohol and substance use, dietary intake, and medication use. Clinical and demographic variables, including age, height, weight, body mass index (BMI), HIV infection status, tobacco use during pregnancy, gestational age, maternal education, occupational status, marital status, and residential area were recorded.

The Alcohol Use Disorders Identification Test (AUDIT) (Babor et al., 2001), an internationally (Bohn et al., 1995) and locally (Nadkarni et al., 2019) validated 10-item self-report screening tool for harmful and HAU patterns, was used as the primary measure of alcohol consumption during pregnancy. Although self-reported prenatal alcohol use is susceptible to stigma-associated underreporting, evidence of the strong self-report measure and biomarker concordance in our study population supports the reliability and accuracy of the reporting (May et al., 2018), and supports its use in the study. AUDIT scores, which can range between 0 and 40, were assessed as both continuous and binary measures. An AUDIT score of eight or more typically defines a higher risk of alcohol-related health issues in non-pregnant, otherwise healthy individuals (Babor et al., 2001), while scores of zero indicate abstinence, and scores between these values reflect risky alcohol use in the context of pregnancy. As an AUDIT score threshold of ≥ 7 has been increasingly applied to account for the increased physiological vulnerability of females to alcohol-related harm (Nadkarni et al., 2019; Parker et al., 2024; Stockton et al., 2025), participants scoring ≥ 7 in our study were considered to have consumed alcohol hazardously during their pregnancy and were included in the HAU group. Those with scores below the threshold were included in the non-hazardous alcohol use (nHAU) group. Data on the quantity, frequency, and gestational timing of alcohol exposure (Sobell and Sobell, 1992; May et al., 2013) were recorded for all participants, and physiological biomarkers of alcohol use, namely phosphatidylethanol (PEth) and ethyl glucuronide (EtG), were measured from blood spots and fingernail clippings in a subset to validate self-report accuracy (May et al., 2018, p. 201; Hasken et al., 2023). Further details on the maternal questionnaire and alcohol use assessment methodology are available in parent study publications elsewhere (May et al., 2018; Hasken et al., 2023).

Information relevant for analysis of the biological samples was collected alongside each sample. Stool consistency, which reflects water content and activity in the colon and is correlated with transit time, was assessed using the Bristol Stool Scale (BSS) (Vandeputte et al., 2016). These scores were categorised into “hard”, “normal” and “loose” stool (Blake et al., 2016), and included as a covariate in fecal analysis. Antibiotic use, known to significantly impact gut microbiota composition and structure, was also noted.

### FASD diagnosis

Kalberg *et al*. (2019) found that assessment of a combination of growth, dysmorphology, and neurobehavioral characteristics in the current cohort enables accurate identification of most children with FASD as early as nine to 18 months of age (Kalberg et al., 2019). As such, an active case-ascertainment procedure was used to diagnose FASD in infants by triangulating data from infant dysmorphology examinations, neurodevelopmental assessments, and maternal interview data (Hoyme et al., 2005, 2016). FASD diagnoses were made with reference to the revised Institute of Medicine criteria described by Hoyme *et al.,* (2016), which contains guidelines for diagnosis of four FASD sub-types (in order of increasing severity): alcohol-related birth defects (ARBD), alcohol-related neurodevelopmental disorder (ARND), partial fetal alcohol syndrome (pFAS) and fetal alcohol syndrome (FAS). Infants were placed into two diagnostic categories for the purpose of this study, namely, a FASD and Not FASD group, based on their latest diagnosis. Participants were assigned to the same group as their child. Participants were excluded from the FASD-related fecal and vaginal microbiota analysis if their infant’s diagnosis was deferred pending further assessment or if their child had not been clinically assessed.

### Biological sampling

Participants provided a self-collected stool sample at a single time point during the second or third trimester of their pregnancy. Stool samples were collected using EasySampler Stool Collection kits and PSP Stool Collection Tubes (Invitek Molecular, Birkenfeld, Germany). Participants enrolled in the study were also asked to, where possible, provide a vaginal swab sample on the day of delivery. The vaginal swab samples were collected from the posterior fornix during speculum examination by registered nurses using OmniGene Vaginal Collection Tubes (DNA Genotek, Ottawa, Ontario, Canada). Following collection, the stool and vaginal swab samples were transported to the Neuropsychiatric Genetics Laboratory at the Stellenbosch University Faculty of Medicine and Health Sciences and stored at-20°C prior to deoxyribonucleic acid (DNA) extraction.

### DNA extraction and targeted amplicon sequencing

Microbial DNA was extracted from 1.4 ml of stabilized stool homogenized in Stool DNA Stabilizer using the PSP Spin Stool DNA Plus Kit (Invitek Molecular) according to manufacturer’s instructions. Microbial DNA was extracted from the vaginal swab samples using the Omega Bio-tek E.Z.N.A. Universal Pathogen Kit (Omega Bio-tek, Norcross, GA, USA). Each extraction batch included a negative control, an extraction blank of Stool DNA Stabilizer (Invitek Molecular), and a positive control, ZymoBIOMICS Microbial Community Standard (Zymo Research, Irvine, CA, USA) prepared according to manufacturer’s specifications.

DNA quantity and quality was evaluated using a Nanodrop One Microvolume Ultraviolet-Visible spectrophotometer (ThermoFisher Scientific, Waltham, MA, USA), and DNA quantity was evaluated using a Qubit 4 Fluorometer (Invitrogen, ThermoFisher Scientific, Waltham, MA, USA) using the double stranded DNA Broad Range assay (Invitrogen). Guided by Qubit concentrations, samples were diluted to 25.0 ng/µl, where possible, using a Hamilton STARlet (Hamilton, Reno, NV, USA). Samples with concentrations below 25.0 ng/µl were included at their measured concentration and flagged in downstream analysis.

Sequencing-ready libraries were generated using a modified Illumina 16S metagenomic protocol and sequenced by the Centre for Proteomic and Genomic Research (CPGR, Cape Town, South Africa). The V1 - V2 hypervariable region of the 16S ribosomal RNA (rRNA) gene was amplified using a 27F/338R primer pair (Alcon-Giner et al., 2017), and unique dual indices and Illumina sequencing adapters were attached to the amplicons using the Nextera XT Index kit (Illumina, San Diego, CA, USA). A positive control (ZymoBIOMICs Microbial Community DNA standard) was included in the final library pool to ensure that the prepared library matched size and composition expectations. The 5 pM pooled sequencing libraries were combined with the PhiX control (Illumina) at a spike-in concentration of 10% v/v.

Denatured fecal microbial DNA library pools were sequenced on an Illumina MiSeq sequencing instrument using the Illumina MiSeq Reagent version 3 kit (600 cycles) programmed to perform a paired-end, dual indexed 2 x 301 cycle sequencing run. The denatured vaginal microbial DNA library pool was sequenced on an Illumina MiSeq sequencing instrument using the Illumina MiSeq Reagent version 2 Nano Kit (500 cycles) programmed to perform a paired-end, dual indexed 2 x 251 cycle sequencing run. Base calling was performed and FASTQ files were generated using Illumina Casava Pipeline 1.8.2. Initial quality assessment was based on data passing the Illumina Chastity filter.

### Cohort descriptive statistics

Case control and other descriptive analyses were conducted using R (version 4.4.1). A Shapiro-Wilk normality test was performed to assess data distribution. The HAU and infant FASD diagnosis group profile variables were summarized using standard summary statistics and represented as means and standard deviations (SD) if normally distributed, medians and interquartile range (IQR) if non-normally distributed, and counts and percentages for categorical data. Differences between groups in normally and non-normally distributed data were assessed using Student’s T-tests and Wilcoxon-Rank Sum tests, respectively. Chi-squared tests and Fisher exact tests (for variables with fewer than five observations for a group) were used to assess differences between groups for categorical variables. A *p*-value < 0.05 was considered statistically significant, while *p*-values < 0.10 were considered nominally significant.

### Bioinformatic analysis

#### Detailed methods are provided in the Supporting Information

Raw paired-end sequence reads were filtered using the Divisive Amplicon Denoising Algorithm 2 (*DADA2*) v.1.32.0 (Callahan et al., 2016) package. Filtered reads were further dereplicated, denoised, and merged using default *DADA2* parameters. Taxonomic assignment was performed using the *DADA2*-formatted SILVA SSU rRNA reference database v.138.2 (Quast et al., 2013; Callahan et al., 2016).

For the fecal microbiota analyses, a feature table comprising of 41,591 amplicon sequence variants (ASVs) derived from 207 fecal samples, with an average read length of ∼ 356 bp, was constructed. For the vaginal microbiota analysis, a second independent feature table consisting of 3,846 ASVs across 28 samples, with read lengths averaging ∼ 351 bp, was created. The ASV tables were separately merged into a unified object with their corresponding taxonomic classifications and relevant participant metadata using *phyloseq* v.1.48.0 (McMurdie and Holmes, 2013). Sequences that remained unassigned or were not classified as “Bacteria” at kingdom-level, and those receiving “Chloroplast” or “Mitochondria” classifications at order and family level, respectively, were removed. Following filtering, 41,485 ASVs in the fecal sample dataset and 2,968 ASVs in the vaginal swab sample dataset were retained for downstream analysis. Taxa were agglomerated and investigated at the genus-level rank.

Rarefaction curves were generated using the *rarecurve* function of *vegan* v.2.6-10 (Dixon, 2003) to determine whether the minimum sequencing depth across the samples adequately captured the bacterial diversity within each dataset. The minimum read depth was found to be adequately representative of diversity, as evidenced by the plateauing of the rarefaction curves across the majority of the samples (**Fig. S1**). The datasets were normalised by random sub-sampling without replacement using the *rarefy* function of the *phyloseq* (McMurdie and Holmes, 2013) package to an even depth of 18,665 and 27,331 reads for the fecal microbiota alcohol use and FASD-related analyses, and 60,667 and 81,236 reads for the vaginal microbiota alcohol use and FASD-related analyses, respectively.

Covariate adjustment sets were selected for each analysis based on their independent association/s with alpha-and beta-diversity measures, statistically significant differences of variables between groups (*p* ≤ 0.05) and their biological relevance. Metadata variables with too few observations for reliable statistical analysis in datasets with limited sample sizes were excluded from models. Linear regression models and permutational ANOVAs (PERMANOVAs), with 999 permutations, were used to test covariate associations with alpha-diversity metrics and beta-diversity distance matrices, and derive *p*-values and effect sizes, respectively. Covariates were retained for adjustment in downstream analyses if significantly associated with a diversity metric (p ≤ 0.05) or if they explained ≥ 2% of variance in beta diversity (R^2^ ≥ 0.02). Covariate adjustment sets are detailed in **Table S1**.

Alpha and beta microbial diversity measures were generated using *vegan* (Dixon, 2003). Alpha diversity was assessed using observed richness, Shannon, and inverse Simpson diversity metrics. Differences in alpha diversity metrics between groups were assessed using robust linear regression models, accounting for relevant covariate adjustment sets. Bray-Curtis dissimilarity matrices were constructed and non-metric multi-dimensional scaling (NMDS) plots were generated to visualize the microbial community structure. PERMANOVAs (9,999 permutations) were performed to determine whether variables of interest were associated with differences in microbial community composition.

*PICRUSt2* (v.2.6.1) (Douglas et al., 2020) was applied to the ASV sequences of the non-rarefied gut microbiota dataset to predict gene family abundances and the related functional pathways of the bacterial communities present. KEGG ortholog (KO) abundances were converted to KEGG pathway abundances (https://www.genome.jp/kegg/) using *ggpicrust2* v.2.5.10 (Yang et al., 2023), and MetaCyc pathway abundances were directly obtained from the *PICRUSt2* pathway inference module. Functional analysis was only performed for the fecal microbiota analysis, as functional inference tools have demonstrated poor performance when applied to vaginal samples (Carter et al., 2023).

MaAsLin3 (v.0.99.16) (Nickols et al., 2026) was used to test associations between metadata variables and microbial community features. Analyses were run on non-rarefied data using default package parameters, with a minimum prevalence threshold of 10% and a minimum relative abundance of 0.001. In addition to the relevant covariate set for each analysis, extraction batch was modelled as a random effect and read depth was included as a covariate to account for differential sequencing depth across samples. Differential taxonomic and functional abundance analyses used a two-sided test of hypothesis, a significance level of *p* ≤ 0.05, and an adjusted *p-*value following Benjamini-Hochberg correction for multiple testing termed *q*. A modest significance threshold *q* ≤ 0.10, chosen to enable a broader view of associations, was used. Genera with a nominal *p*-joint ≤ 0.05 were reported on in the absence of false discovery rate (FDR)-significant results for the purposes of this exploratory study.

## Results

### Study cohort profile

Of the total cohort (n = 213) recruited, 207 participants provided stool samples and met criteria for inclusion in the primary aim of the study. These participant characteristics are described in **Table 1**. The median age and BMI of the participants was 26 years (IQR: 22 – 32) and 24.4 kg/m^2^ (IQR: 21.0 - 29.7), respectively. Of these participants, 138/207 (66.7%) consumed alcohol hazardously during their pregnancy (HAU: AUDIT ≥ 7), while 69/207 (33.3%) consumed little-to-no alcohol during pregnancy (nHAU: AUDIT score < 7). Fecal samples were collected at a median gestational age of 20 weeks (IQR: 14 - 26) and no significant difference in sampling timepoint was identified between groups (*p* = 0.45).

**Table 1:**
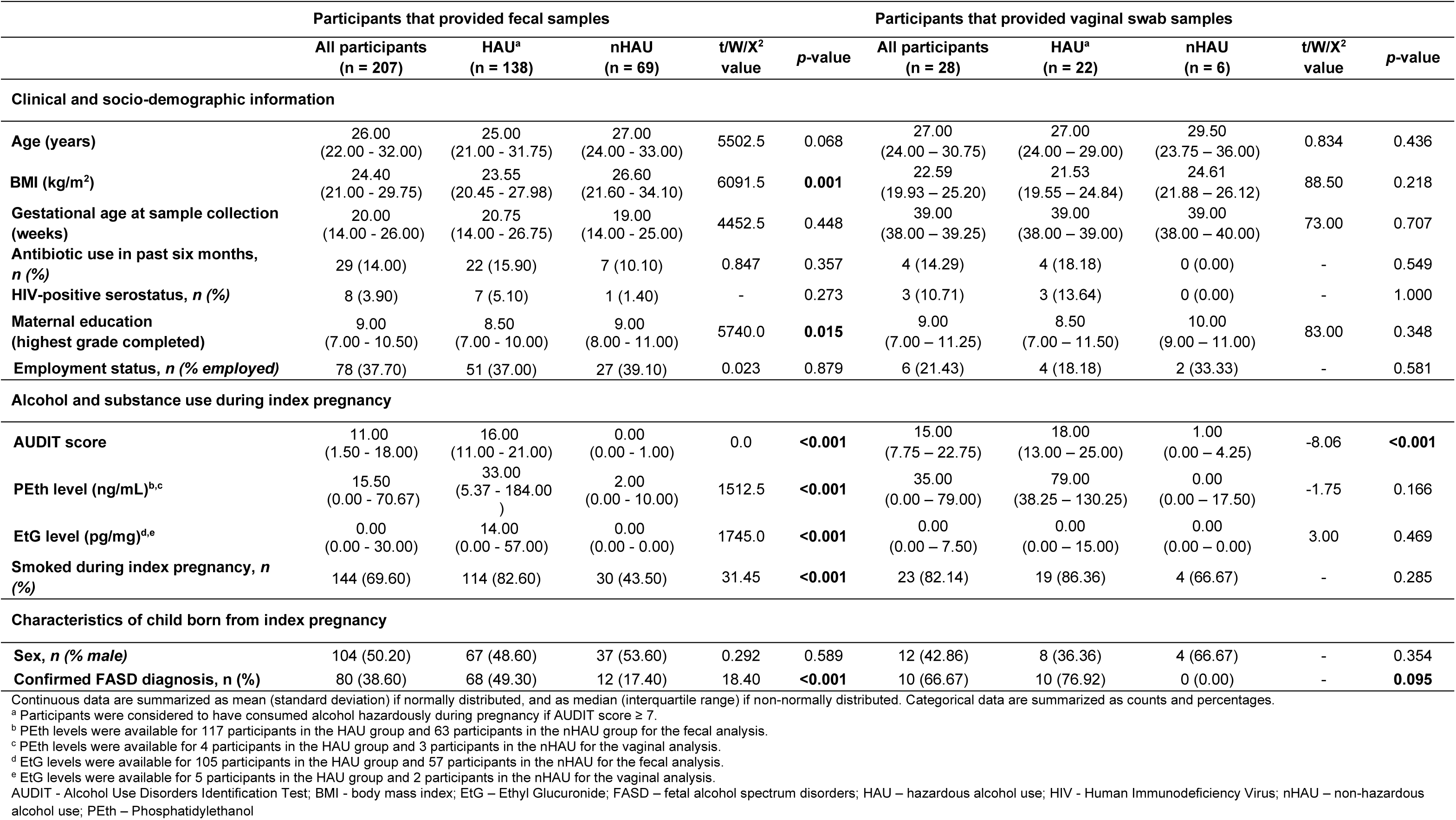
Characteristics of the study participants included in the cross-sectional fecal (n = 207) and vaginal microbiota (n = 28) alcohol-use investigation.

Participants in the HAU group had a lower median BMI and level of educational attainment, and significantly higher median self-reported and biological measures of alcohol use compared to the nHAU group (*p* < 0.05). A greater proportion of those reporting HAU also reported smoking during the index pregnancy (*p* < 0.001).

A subset of participants that provided stool samples provided a vaginal swab sample on the day of birth (n = 28/207; 13.5%). This sample cohort displayed similar characteristics to those in the larger fecal microbiota cohort (**Table 1**). The participants had a median AUDIT score of 15.0 (7.8 - 22.8), with 22/28 (78.6%) having consumed alcohol hazardously during their pregnancy and reporting significantly higher median AUDIT scores relative to the nHAU group (*p* < 0.001).

Of the 207 participants that provided stool samples, 168 (81.2%) met criteria for inclusion in the secondary aim of the study. Participant characteristics are summarized in **Table S2**.

The median age and BMI of the participants was 26 years (IQR: 22 - 27) and 24.9 kg/m^2^ (IQR: 21.1 - 31.0), respectively. Of these participants, 80/168 (47.6%) had an infant with an FASD diagnosis (FASD group) and 88/168 (52.4%) gave birth to an infant without an FASD diagnosis (Not FASD group). Fecal samples were collected at a median gestational age of 19 weeks (IQR: 14 – 26) and no significant differences in sampling timepoint during pregnancy were identified between groups (*p* = 0.26).

Besides poorer maternal educational attainment and higher self-reported and biological measures of alcohol use in the FASD group relative to the Not FASD group, no significant differences in participant characteristics were identified between the groups (*p* > 0.05). The median AUDIT score for the sample was 11.0 (IQR: 0.0 – 17.0). HAU was reported in 85.0% of participants in the FASD group compared to 45.5% in the Not FASD group (*p* < 0.001).

Of the vaginal swab samples collected, samples from 13 participants were excluded: one due to infant death, six due to Caesarean-section delivery, two due to geographical relocation prior to infant assessment, and four due to absence of confirmed diagnoses. The 15 remaining participants had a mean AUDIT score of 14.32 (SD = 8.70), with 13 (86.7%) of these participants having consumed alcohol hazardously during their pregnancy. Of these participants, 10/15 (66.7%) participants gave birth to an infant who received an FASD diagnosis and 5/15 (33.3%) gave birth to infants without FASD.

### Maternal fecal microbiota phylum-and genus-level composition

The most abundant phyla across all the fecal samples were *Bacillota, Bacteroidota, Pseudomonadota, Actinomycetota, Thermodesulfobacteriota, Verrucomicrobiota and Cyanobacteriota* (**Table S3**).

At the genus-level, the fecal microbiota (primary aim) was dominated by *Segatella* (formally *Prevotella*), which was present at a median relative abundance of 23.3% (**Fig. 1A, Table S4**). In addition, and listed in order of decreasing median abundance, *Faecalibacterium* (4.4%), *Succinivibrio* (3.9%), *Oscillospiraceae UCG-002* (3.6%), *Bacteroides* (3.1%), *Dialister* (1.9%), *Leyella* (1.5%), *Lachnospiraceae NK4A136* group (1.2%), *Roseburia* (1.1%) and *Sutterella* (1.1%), were among the remaining top ten most abundant bacterial genera present in the samples (**Fig. 1A, Table S4**). Similar proportions were identified among the 168 participants who provided fecal samples and had an infant who had undergone diagnostic assessment for FASD (secondary aim) **(Fig. 1B, Table S4)**.

**Figure 1:**
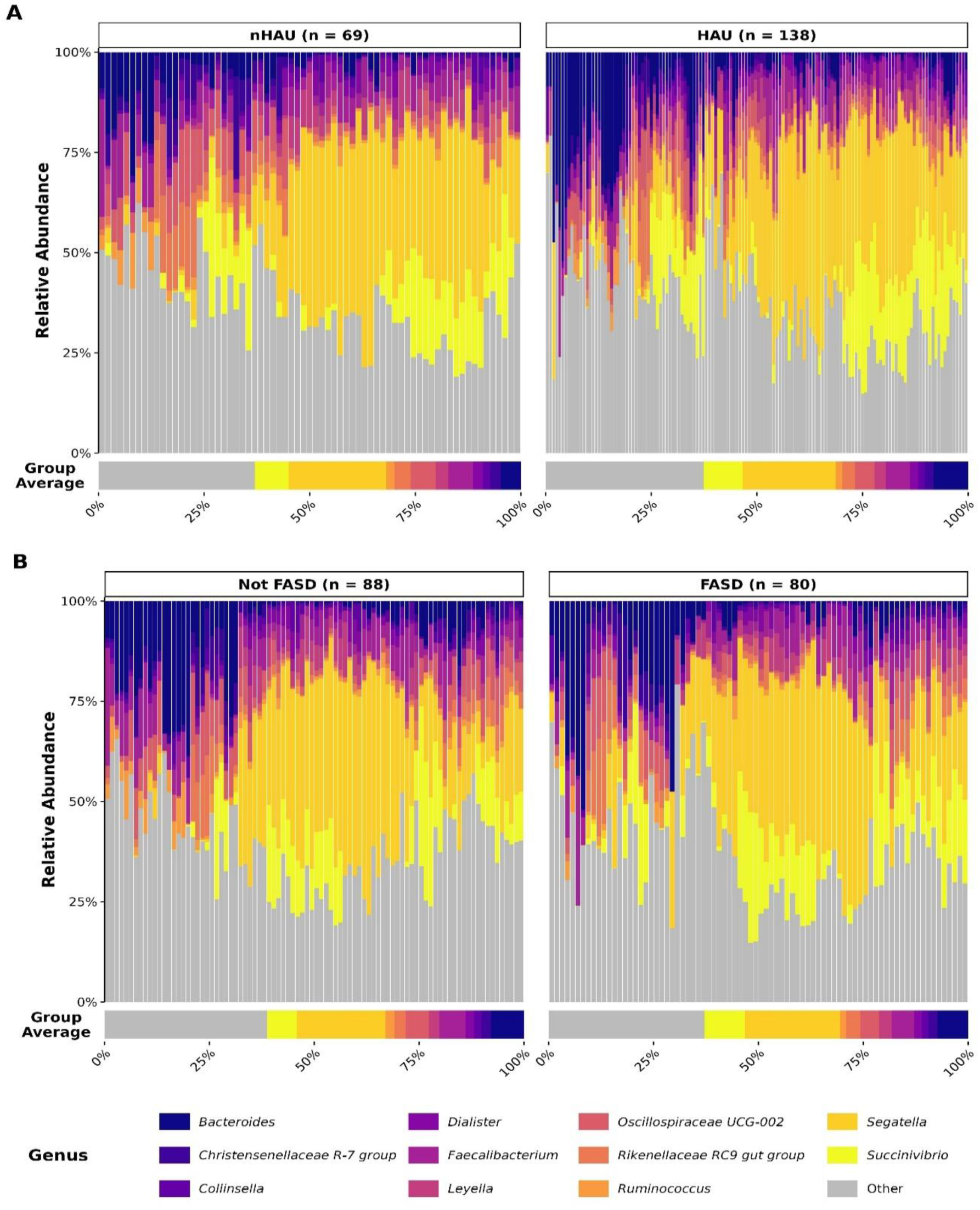
Genus-level composition of the maternal fecal microbiota across alcohol use and infant FASD diagnostic groups. Stacked bar charts display the top ten most abundant bacterial genera present in the maternal fecal samples, with samples ordered by Bray-Curtis dissimilarity and facetted by (**A**) HAU (AUDIT ≥ 7: n = 138; AUDIT < 7: n = 69) and (**B**) infant FASD diagnostic category (FASD: n = 80; Not FASD: n = 88). Horizontal bar charts below each panel display the mean relative abundance per facetted group. AUDIT – Alcohol Use Disorders Identification Test; FASD – fetal alcohol spectrum disorders; HAU – hazardous alcohol use.

### Maternal fecal microbiota diversity measures relative to alcohol use and infant FASD diagnosis

Analyses revealed that AUDIT score was negatively associated with alpha diversity measures (observed richness, Shannon and Inverse Simpson measures; *p* < 0.05; Fig. **2A**). Observed richness did not differ according to HAU group (*p* = 0.09; **Fig. 2B**) but was significantly lower in the FASD compared to the Not FASD group (*p* = 0.046; **Fig. 2C**). No significant associations were identified for the remaining diversity indices (*p* > 0.10).

**Figure 2:**
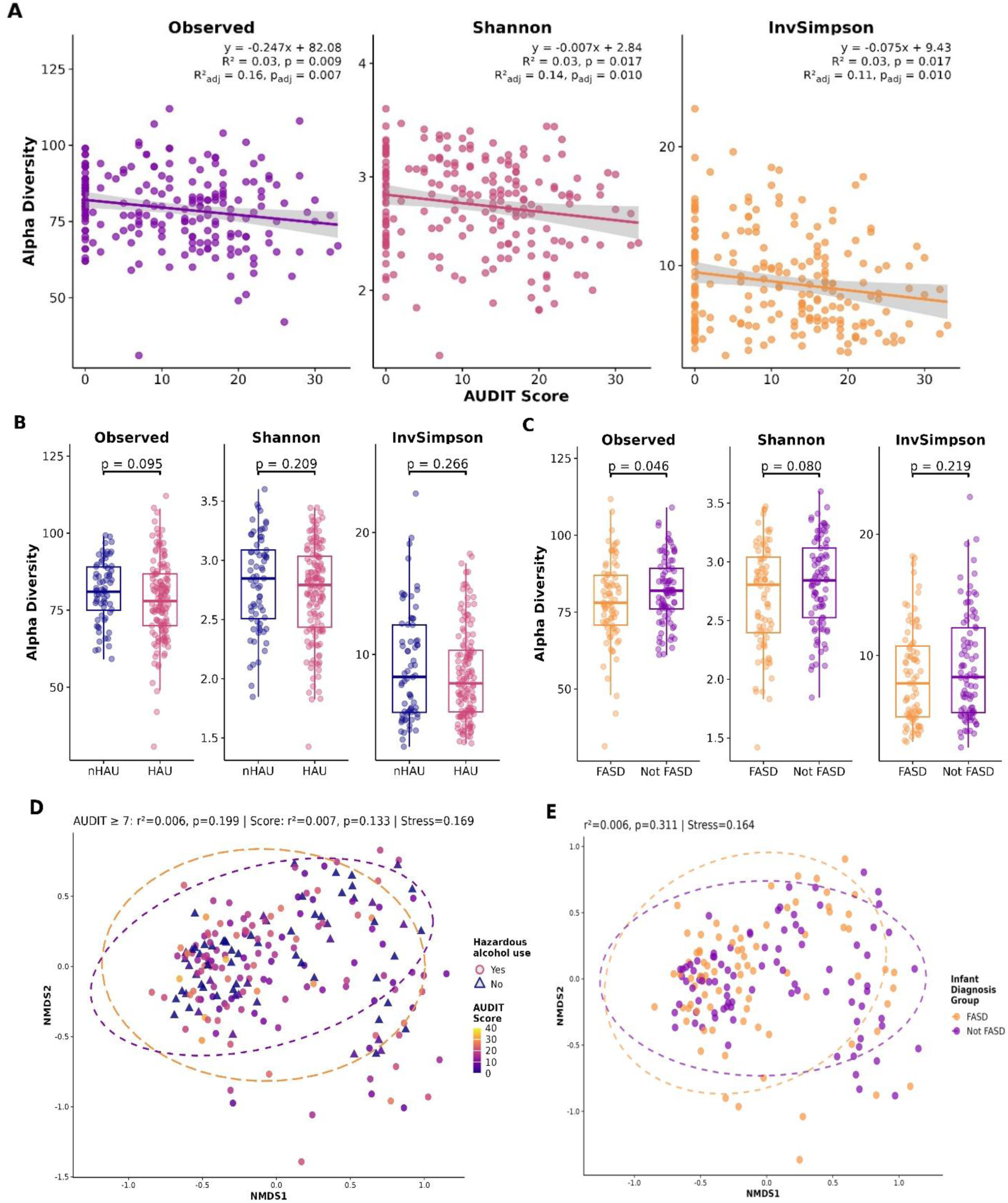
Maternal fecal microbiota diversity relative to AUDIT score and infant FASD diagnosis. Alpha-and beta-diversity analyses were conducted in the full maternal cohort (n = 207) for alcohol use measures and in a subset with available infant FASD diagnostic data (n = 168). Linear regression and box-and-whisker plots of alpha-diversity indices (Observed richness, Shannon and Inverse Simpson) are shown in relation to (**A**) continuous AUDIT score (n = 207), (**B**) dichotomized AUDIT score reflecting HAU during pregnancy (AUDIT ≥ 7: n = 138; AUDIT < 7: n = 69) and (**C**) infant FASD diagnostic group (FASD: n = 80; Not FASD: n = 88). NMDS Bray-Curtis dissimilarity ordination plots are shown for (**D**) investigated alcohol use measures, colored by AUDIT score and stratified by alcohol use group, and (**E**) infant FASD diagnostic group. AUDIT – Alcohol Use Disorders Identification Test; FASD – fetal alcohol spectrum disorders; HAU – hazardous alcohol use; NMDS – non-metric multidimensional scaling.

No significant differences in beta diversity were identified for any of the outcomes investigated (*p* > 0.05, PERMANOVA, **Fig. 2D**, **Fig. 2E**).

### Differentially abundant maternal fecal microbiota associated with alcohol use and infant FASD

No differentially abundant taxon associations in the MaAsLin3 [69] joint models survived FDR correction (q ≤ 0.10) in either the alcohol use or infant FASD diagnosis analyses. The genera with the strongest joint nominal associations (p < 0.05) for each analysis are presented in **Fig. 3** and **Table S5**, and reported below.

**Figure 3:**
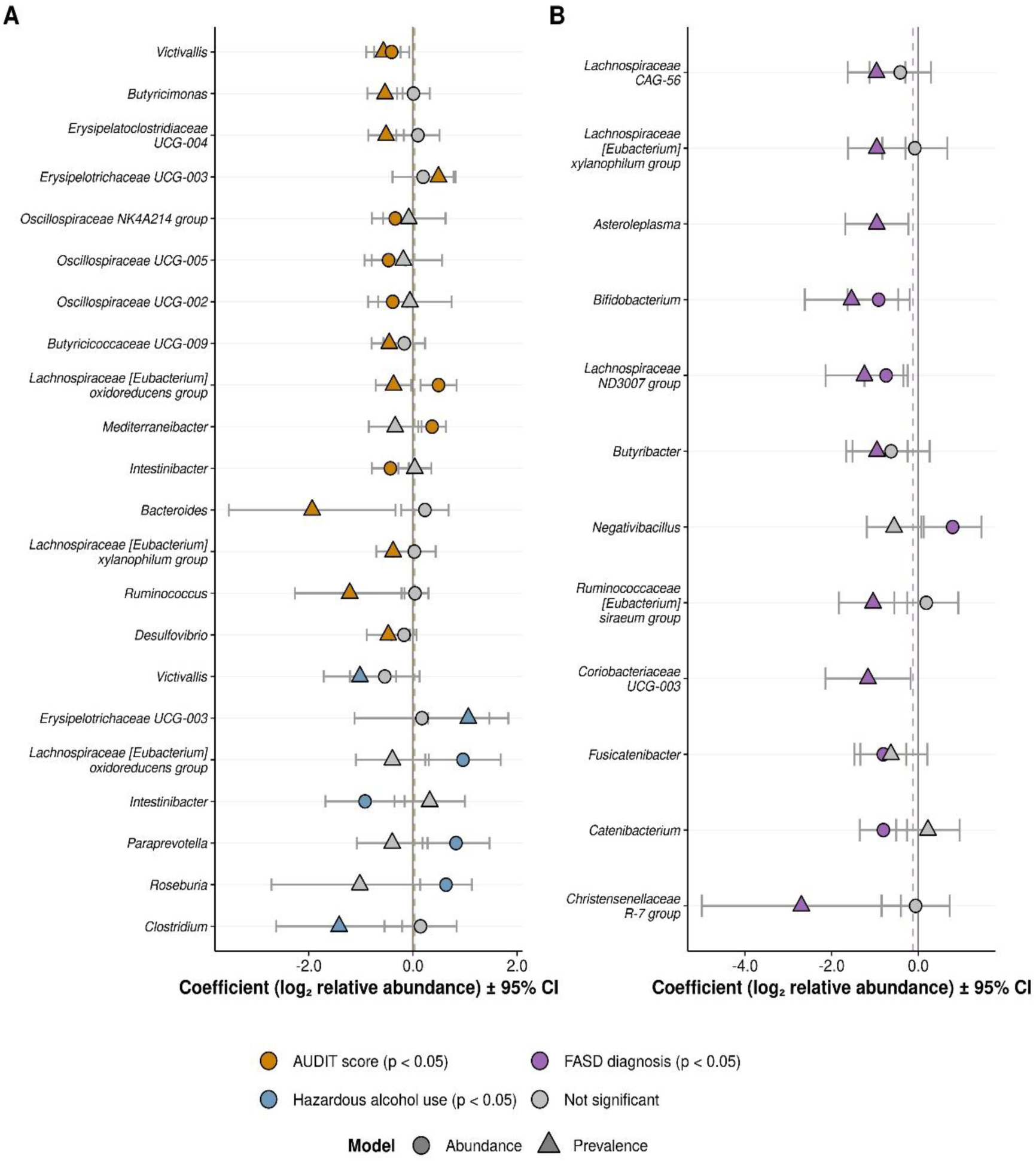
Maternal fecal bacterial genera nominally associated with alcohol use and infant FASD diagnosis. Genera, ordered by the most to least significant joint nominal p-value ≤ 0.05, are presented on the y-axis for (**A**) continuous AUDIT score (orange) and HAU (AUDIT ≥ 7; blue), and (**B**) infant diagnostic category (purple). Each plotted feature contributes two data points; namely, a filled circle representative of the abundance model (log_2_ relative abundance) co-efficient and a filled triangle denoting the prevalence model (log_2_ relative abundance) co-efficient, with their respective 95% confidence intervals. The coefficients of the binary analysis reflect the HAU and FASD group relative to controls. Grey point fill color represents an individual model with a *p* ≥ 0.05. Dashed vertical lines delineate the median null hypothesis coefficient for the abundance models (AUDIT (orange): 0.1, HAU (blue): 0.3, and FASD diagnosis (purple):-0.12) and the solid vertical line marks zero. AUDIT – Alcohol Use Disorders Identification Test; FASD – fetal alcohol spectrum disorders; HAU – hazardous alcohol use.

Among the nominally significant genera, 15 displayed suggestive associations with AUDIT score (**Fig. 3A**). Of these, 14 genera had concordant abundance and prevalence co-efficient directions, of which two were positively and 12 were negatively associated with higher AUDIT scores. *Victivallis* showed the strongest nominal association, with lower relative abundance and prevalence observed in participants with higher scores (*p*-joint = 0.002, coef_abun_=-0.41, *p*_abun_ = 0.02; coef_prev_=-0.57, p_prev_=0.001). The *Lachnospiraceae [Eubacterium] oxidoreducens* group showed a nominal joint association (*p-*joint = 0.02), but presented discordant abundance and prevalence co-efficient directions (coef_abun_ = 0.49, coef_prev_ =-0.37). Additionally, three *Oscillospiraceae* genera displayed negative nominal associations with AUDIT score primarily driven by abundance models.

Seven genera were nominally associated with HAU during pregnancy (p < 0.05; **Fig. 3A**). Similar to AUDIT score, *Victivallis* showed the strongest nominal association (*p-*joint = 0.008), driven by lower prevalence observed in the HAU group relative to the NHAU group (coef_prev_=-1.02, *p*_prev_=0.004). *Erysipelotrichaceae UCG-003*, the *Lachnospiraceae [Eubacterium] oxidoreducens* group and *Intestinibacter* were also nominally associated with HAU, with coefficient directions and *p*-values consistent with those observed with AUDIT score.

Twelve bacterial genera were nominally associated with the FASD group (p < 0.05). The majority of the implicated genera displayed lower relative abundance and prevalence in the FASD group. Of these, ten genera displayed concordant co-efficient directions across both models, all in the negative direction. Reductions in *Bifidobacterium* and *Lachnospiraceae ND3007* group in the FASD group were supported by nominal associations in both models (*Bifidobacterium* coef_abun_ = −0.91, *p*_abun_ = 0.04; coef_prev_ = −1.54, p_prev_ = 0.01; *p*-joint = 0.01 and *Lachnospiraceae ND3007* group coef_abun_ = −0.74, *p*_abun_ = 0.02; coef_prev_ = −1.24, *p*_prev_ = 0.01; *p*-joint = 0.01; **Fig. 3B**).

### Predicted maternal fecal microbiota functional potential relative to alcohol use and infant FASD

No individual KEGG pathways reached significance in the AUDIT score, HAU and infant FASD diagnosis analysis following multiple test correction *(q* > 0.10, **Fig. 4**, **Table S6**).

**Figure 4:**
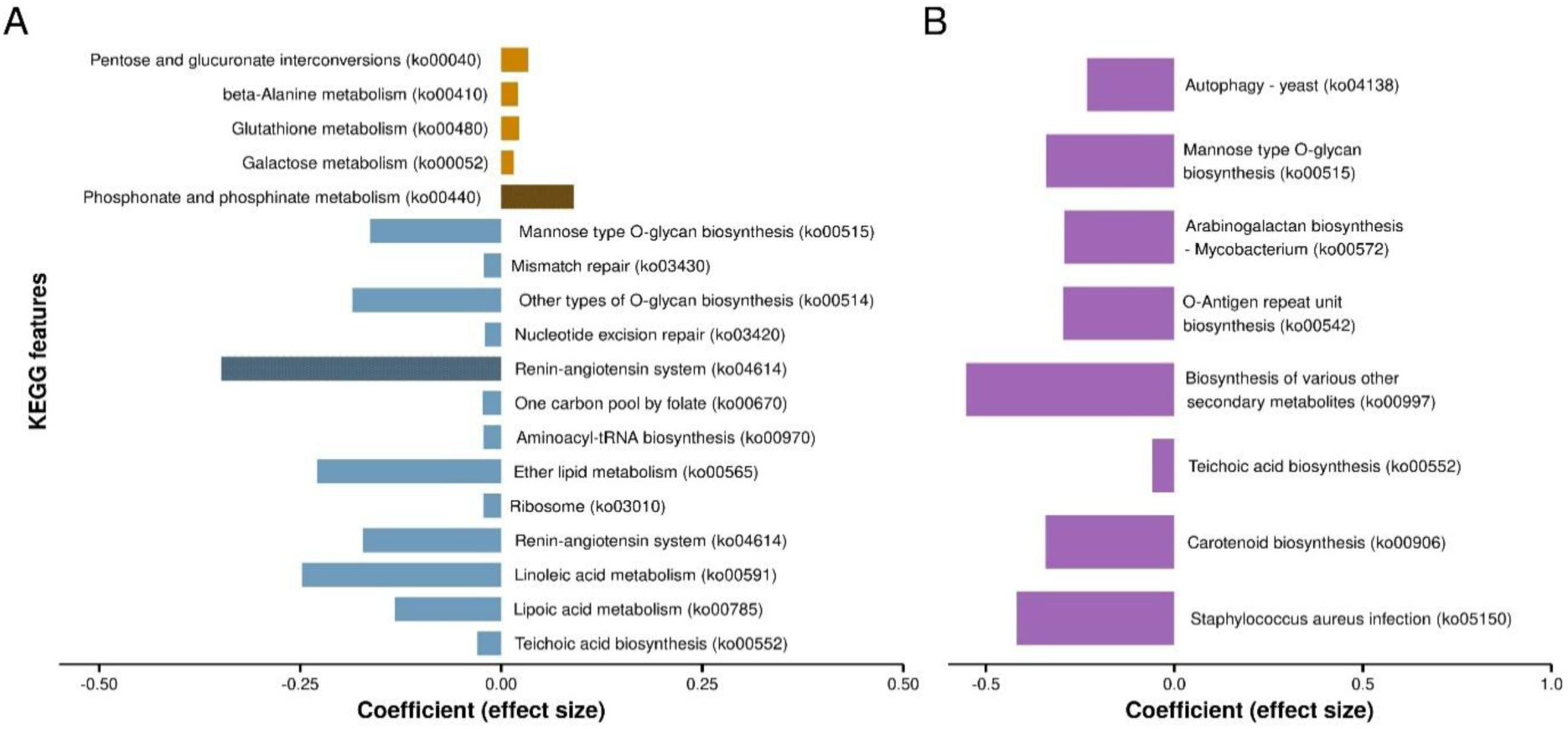
KEGG pathways nominally associated with alcohol use and infant FASD diagnosis in maternal fecal samples. Horizontal bar plots display the coefficients (effect sizes) of the KEGG features nominally associated (*p* < 0.05) with (**A**) AUDIT score and HAU (AUDIT ≥ 7; darker cross-hatched bars) and (**B**) infant FASD diagnostic category. Bars extending to the right indicate positive associations (enrichment), and bars extending to the left represent negative associations (depletion), with the measure or group investigated. Features are grouped by direction and are ordered by *p*-value within each group, with the most significant features appearing at the extremes of the plot for each group (top rank for enriched, bottom rank for depleted). Unannotated BRITE hierarchy entries were excluded from the figures. AUDIT – Alcohol Use Disorders Identification Test; FASD – fetal alcohol spectrum disorders; HAU - hazardous alcohol use; KEGG – Kyoto Encyclopedia of Genes and Genomes.

Twenty-three nominally significant associations were observed in the AUDIT score analysis (*p* < 0.05), with the majority (15/23; 65.2%) displaying reduced predicted functional capacity with increasing score (**Fig. 4A**). The strongest nominal associations were identified in pathways related to metabolism and genetic information processing, including teichoic acid biosynthesis (*p* = 0.007), pentose and glucuronate interconversions (*p* = 0.019), lipoic acid metabolism (*p* = 0.027), ribosome (*p* = 0.033) and nucleotide excision repair (*p* = 0.041). For HAU, two pathways displayed nominally significant associations (*p* < 0.05): Renin-angiotensin system (*p* = 0.040) and phosphonate and phosphinate metabolism (*p* = 0.048, **Fig. 4A**).

All eight annotated KEGG pathways displayed negative nominal associations (*p* < 0.05) in the FASD group (**Fig. 4B**). The strongest of these associations was *Staphylococcus aureus* infection (*p* = 0.003), followed by several cell structure biosynthesis pathways, including teichoic acid biosynthesis (*p* = 0.016), O-Antigen repeat unit biosynthesis (*p* = 0.029), arabinogalactan biosynthesis (*p* = 0.030) and mannose type O-glycan biosynthesis (*p* = 0.036).

AUDIT score was nominally associated with ten MetaCyc pathways (*p* < 0.05; **Fig. 5A, Table S7**). The strongest nominal association involved a reduction in phospholipases (β = −0.349, *p* = 0.003), a fatty acid and lipid degradation metabolism pathway. Nitrogen and amino acid metabolism pathways, including L-arginine degradation XIII (*p* = 0.005), the urea cycle (p = 0.021), polyamine biosynthesis II (*p* = 0.008), and peptidoglycan biosynthesis pathways specific to Gram-positive bacteria, *Enterococcus faecium* (*p* = 0.016) and *Staphylococci* (*p* = 0.036), were negatively nominally associated with AUDIT score. Two pathways related to vitamin B1 and B6 biosynthesis were found to be nominally positively associated with AUDIT score.

**Figure 5:**
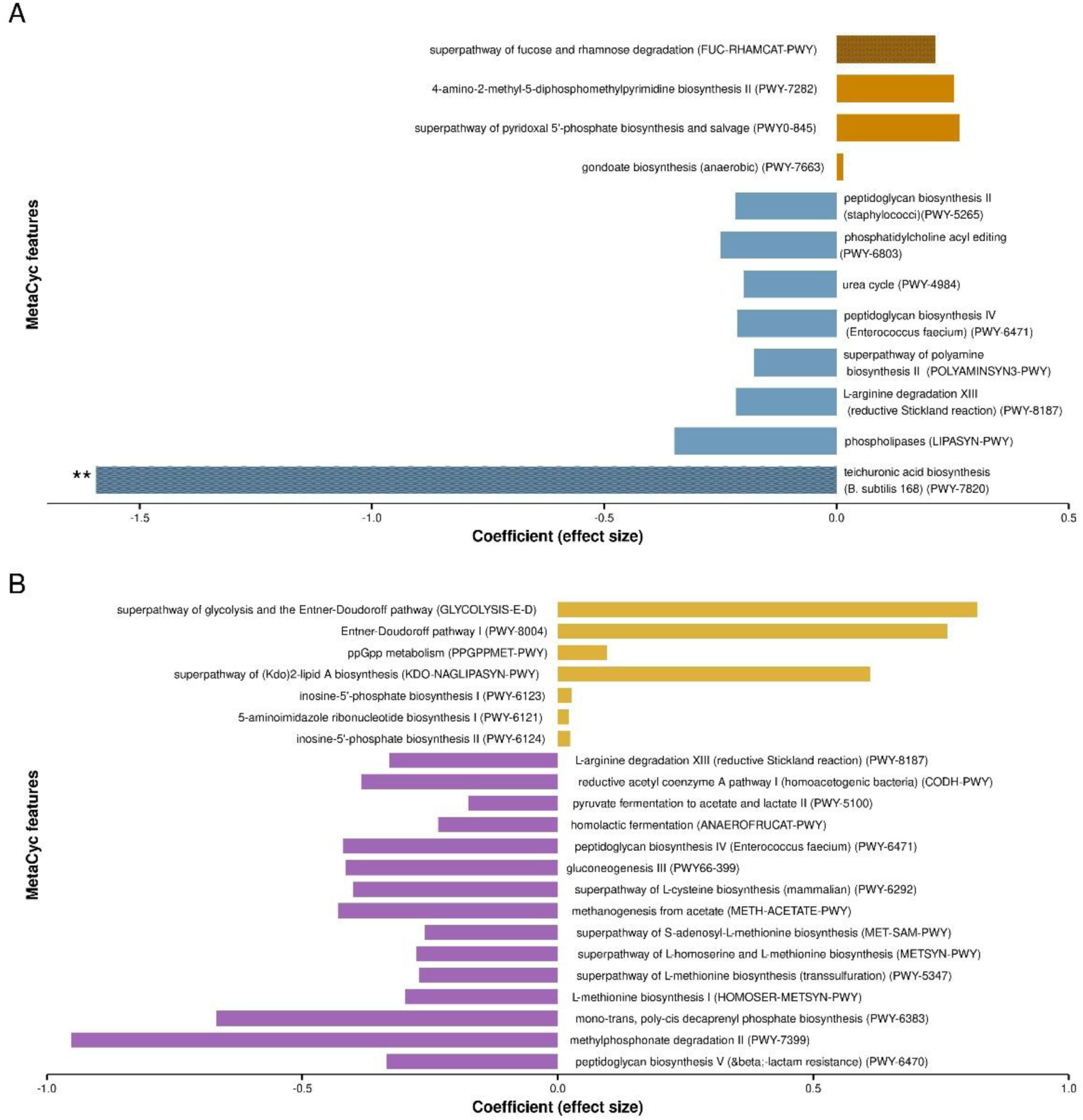
MetaCyc pathways nominally associated with alcohol use and infant FASD diagnosis in maternal fecal samples. Horizontal bar plots display the coefficients (effect sizes) of the MetaCyc features nominally associated (*p* < 0.05) with (**A**) AUDIT score and HAU (AUDIT ≥ 7; darker cross-hatched bars) and (**B**) infant FASD diagnosis. Bars extending to the right indicate positive associations (enrichment), and bars extending to the left represent negative associations (depletion), with the measure or group investigated. Features are grouped by direction and are ordered by *p*-value within each group, with the most significant features appearing at the extremes of the plot for each group (top rank for enriched, bottom rank for depleted). Asterisks denote features significantly associated with outcome of interest (* *q* < 0.10; ** *q* < 0.05; *** *q* < 0.01). AUDIT – Alcohol Use Disorders Identification Test; FASD – fetal alcohol spectrum disorders, HAU – hazardous alcohol use.

The teichuronic acid biosynthesis (*Bacillus subtilis* 168) pathway was significantly reduced in the HAU group (*q* = 0.043), while fucose and rhamnose degradation showed nominally increased predicted abundance (*p* = 0.023, **Fig. 5A, Table S7**).

Twenty-two MetaCyc pathways displayed nominally significant differences between infant FASD diagnosis groups (p < 0.05, **Fig. 5B**, **Table S7**). The predicted functional potential of two pathways approached near-significance (*q* ≤ 0.10), including peptidoglycan biosynthesis V (β-lactam resistance; β =-0.335, *q* = 0.104) and methylphosphonate degradation II (β =-0.953, *q* = 0.110). Of note, six related pathways involved in one-carbon and sulphur amino acid metabolism were nominally reduced in the FASD group, including methylphosphonate degradation II, L-methionine biosynthesis I (*p* = 0.015) and superpathways of L-methionine biosynthesis (transsulfuration; *p* = 0.015), L-homoserine and L-methionine biosynthesis (*p* = 0.018), S-adenosyl-L-methionine biosynthesis (*p* = 0.018), and L-cysteine biosynthesis (mammalian; *p* = 0.021). The predicted abundance of several cell wall biosynthesis pathways was also nominally decreased. Energy metabolism pathways associated with lactic acid fermentation (homolactic fermentation; pyruvate fermentation to acetate and lactate II) displayed nominal reductions in predicted functional potential in the FASD group, while three pathways characteristic of Gram-negative bacterial metabolism, including Entner-Doudoroff pathways and (Kdo)-lipid A biosynthesis, were increased.

### Maternal vaginal microbiota composition, diversity and association with measures of interest

The most abundant bacterial phyla present across the maternal vaginal swab samples, in order of decreasing abundance, were: *Bacillota*, *Actinomycetota*, *Bacteroidota*, *Fusobacteriota* and *Pseudomonadota* (**Table S8**).

At the genus-level, the vaginal bacteriome (primary aim) of the 28 participants was dominated by *Lactobacillus* species, present at a median relative abundance of 72.5% (**Fig. 6A**, **Table S9**). In addition, and listed in order of decreasing median abundance, *Gardnerella* (5.3%), *Prevotella* (0.6%), *Ureaplasma* (0.3%) and *Hoylesella* (0.2%) were among the remaining top five most abundant bacterial genera present in the samples. *Lactobacillus*, *Gardnerella* and *Prevotella* were similarly ranked as the three most abundant genera in the secondary FASD analysis (n = 15); however, the proportions differed substantially relative to those identified in the full dataset (**Fig. 6B**, **Table S9**).

**Figure 6:**
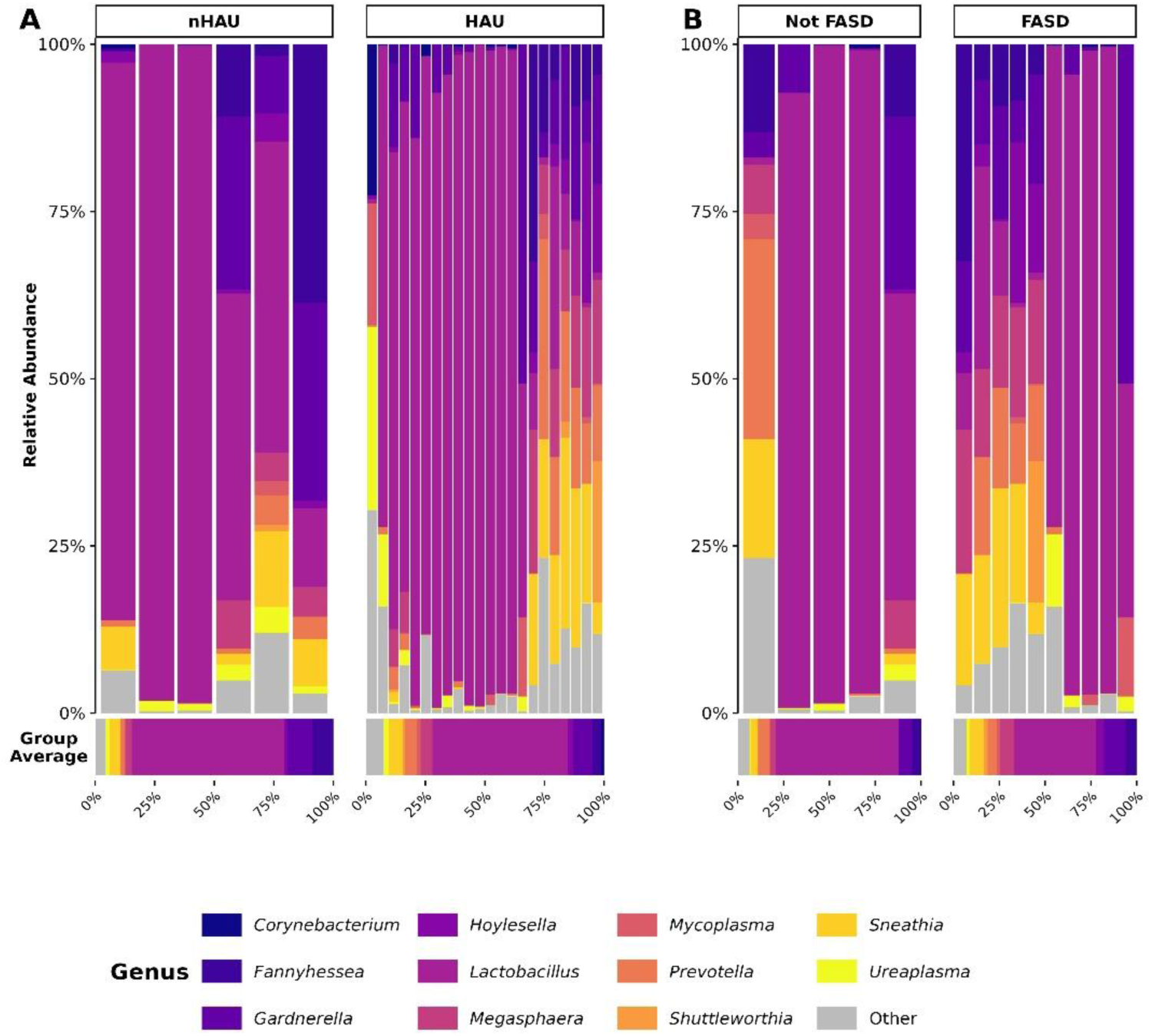
Genus-level composition of the maternal vaginal microbiota across alcohol use and infant FASD diagnostic groups. Stacked bar charts display the top most abundant bacterial genera present in the maternal vaginal swab samples, with samples ordered by Bray-Curtis dissimilarity and facetted by (**A**) HAU (AUDIT ≥ 7: n = 22; AUDIT < 7: n = 6) and (**B**) infant FASD diagnostic category (FASD: n = 10; Not FASD: n = 5). Horizontal bar charts below each panel display the mean relative abundance per facetted group. AUDIT – Alcohol Use Disorders Identification Test; FASD – fetal alcohol spectrum disorders; HAU – hazardous alcohol use.

AUDIT score, HAU or infant FASD diagnostic group were not significantly associated with alpha and beta diversity measures (*p* > 0.05; **Fig. S2**).

For the differential abundance analysis, no genera survived FDR correction in the joint models (q ≤ 0.10) or met the threshold for reporting based on nominal significance (*p*-joint < 0.05) for any analysis.

## Discussion

This study is the first to investigate the maternal gut and vaginal microbiota in relation to prenatal alcohol consumption and infant FASD diagnosis. In a cohort of pregnant South African participants recruited from a community with high FASD prevalence (May et al., 2022), we demonstrate that alcohol use during pregnancy is associated with reduced maternal gut microbiota diversity, with taxonomic and functional differences associated, albeit weakly, with maternal alcohol use and infant FASD diagnosis. In contrast, no associations between maternal vaginal microbiota diversity and composition, alcohol use measures, and infant FASD diagnostic category were found.

Gut microbiota alpha-diversity was negatively associated with AUDIT score, with trend-level reductions in observed richness identified in participants who reported HAU during pregnancy. This is consistent with non-pregnant clinical and population-based studies reporting significantly reduced bacterial alpha-diversity, specifically richness, in individuals with high-risk drinking patterns (Koponen et al., 2025; Liu et al., 2025) and alcohol use disorder (Engen et al., 2015; Piacentino et al., 2024; Sosnowski and Przybyłkowski, 2024), but is in contrast to the higher diversity reported in pregnant alcohol-consuming Chinese participants in a small cross-sectional study (n = 29) (Wang et al., 2021). Evidence from recent rodent studies further supports alcohol-induced gut bacterial diversity reduction in pregnancy (Bodnar et al., 2024; Engen et al., 2015; Sosnowski and Przybyłkowski, 2024). This suggests that alcohol exposure compounds the reduction in bacteria diversity that occurs in response to hormonal and immune changes that support physiological adaptation and fetal health during pregnancy (Nuriel-Ohayon et al., 2016).

Rodent studies have provided evidence of alcohol-induced differences in maternal microbiota composition and metabolomic profiles, which were linked to altered offspring gut colonization and the emergence of neurodevelopmental changes and behaviors resembling FASD (Busayli et al., 2025; Smith, 2025; Virdee et al., 2021). In our study, observed richness was significantly and Shannon diversity nominally reduced in participants who gave birth to infants diagnosed with FASD. The significant reduction in bacterial richness without significant changes in metrics weighted by community evenness in participants with infants diagnosed with FASD suggests the abundance of dominant tax is preserved while that of less abundant or rarer bacterial taxa is lost. As FASD development is determined by alcohol dose, as well as complex interactions among the quantity, frequency, and gestational timing of exposure, maternal and fetal genetics, maternal nutrition, epigenetic modification and concurrent substance use (Popova et al., 2023), we examined infant FASD diagnosis independently from maternal alcohol consumption measures. Whether microbial profile contributes independently to FASD development or reflects the influence of the aforementioned factors requires further investigation.

Gut microbiota community structure was not significantly associated with either self-reported maternal alcohol use or infant FASD diagnosis. Previous findings in non-pregnant cohorts are mixed. Koponen *et al.,* (2025) reported a small but consistent association of alcohol use with weighted UniFrac distances in a study of ∼ 4,600 shallow-shotgun sequenced samples (52.5% female) (Koponen et al., 2025) and Liu *et al.,* (2025) found no significant associations in a predominantly low-income black population (n = 538; 388 females) using Bray-Curtis dissimilarity (Liu et al., 2025). Though Wang *et al*., (2021) found that Jaccard dissimilarity differed significantly between alcohol-consuming and non-alcohol consuming pregnant participants; however, the small sample size, the population group and the dissimilarity metric used limit direct comparison. Our findings also contrast with a recent rodent study that reported community-level differences between alcohol-exposed and control dams (Bodnar et al., 2024), which could reflect species-specific gut microbial community differences and the controlled exposure to alcohol in rats. In the context of infant FASD diagnosis, the absence of significant differences in beta-diversity supports the hypothesis that less abundant bacterial taxa are being selectively lost without significantly altering the overall microbial community structure.

Reductions in the gut relative abundance of several Gram-positive genera involved in intestinal barrier integrity maintenance, including *Bifidobacterium*, *Lachnospiraceae* genera, *Butyribacter* and *Christensenellaceae R-7* group species, were nominally associated with alcohol use and identified in participants whose infants received a diagnosis on the FASD spectrum. These taxonomic shifts are consistent with the predicted reduction of Gram-positive bacterial cell wall biosynthesis pathways, specifically the teichuronic acid biosynthesis (*B. subtilis* 168) pathway with HAU, and the nominally significant reductions in lactic acid fermentation pathways identified in the FASD group. A similar pattern of Gram-positive taxa depletion detected at lower taxonomic levels has been observed in a recent rodent study, with reduced abundances of *Lachnospiraceae* and *Ruminococcus* in alcohol-exposed dams also being noted (Bodnar et al., 2022). Bodnar *et al.,* (2024) found a similar pattern of Gram-positive taxa depletion in *Lachnospiraceae* and *Ruminococcus* detected at lower taxonomic levels in alcohol-exposed rodent dams (Bodnar et al., 2022). In human studies, alcohol-associated enrichment of Gram-negative bacteria and depletion of putatively beneficial Gram-positive taxa, which promote a pro-inflammatory gut microbiome profile, may mediate detrimental downstream health effects via endotoxin release through compromised intestinal epithelial lining (Koponen et al., 2025). Thus, maternal systemic inflammation due to alcohol-induced depletion of the aforementioned SCFA-producing and barrier-maintaining taxa in the gut, combined with the direct effects of alcohol on intestinal permeability, could increase fetal susceptibility to PAE-induced damage. As such, targeted pre-and probiotic intervention aimed at restoring these taxa may represent a promising avenue for mitigating alcohol-associated inflammation and its detrimental impact on the fetus.

Ethanol is a known disruptor of the host one-carbon metabolism (OCM) cycle, primarily exerting its effect through interference with folate absorption and OCM enzyme activity, thereby compromising normal DNA synthesis and methylation processes (Kruman and Fowler, 2014). Our functional analysis also identified nominal reductions in six predicted pathways associated with OCM methionine cycling and transulfuration in the gut microbiota of participants who gave birth to infants with FASD, including those involved in methylphosphonate degradation and L-homoserine, L-methionine, S-adenosyl-L-methionine (SAM) and mammalian L-cysteine biosynthesis. Given fetal reliance on the maternal supply of methyl groups for fetal and placental DNA methylation (McGee et al., 2018), diminished gut microbial contributions to the OCM of participants with infants diagnosed with FASD combined with alcohol-induced maternal OCM dysfunction could directly affect fetal epigenetic programming, which is increasingly recognised as an important mechanism in FASD development (Lussier et al., 2017). In addition, a reduction in the predicted transulfuration pathway capacity, which connects the methionine cycle to antioxidant production through the conversion of homocysteine to cysteine (Chen et al., 2018), points to diminished antioxidant defence in response to alcohol-induced oxidative stress. Though these mechanisms provide plausible explanation for how the maternal gut microbiota contribute to FASD pathogenesis, more evidence is required to validate our findings and support these results.

Vaginal microbiota diversity and composition were not associated with alcohol use or infant FASD diagnostic group. The absence of significant findings may be attributed to the limited diversity of this body site, combined with pregnancy-associated reductions in community diversity and complexity (Nuriel-Ohayon et al., 2016), and the small sub-sample used for the vaginal analysis. Nevertheless, this novel analysis informs study design and sample size considerations for future studies.

The results of this exploratory study should be interpreted within the context of a few important methodological considerations. The cross-sectional design provides single snapshots of the maternal gut and vaginal bacteriome at discrete time points, restricting our ability to simultaneously assess alcohol’s impact on both body sites, delineate temporal changes across pregnancy, and relate these dynamics to infant FASD development risk. As vaginal delivery facilitates the vertical transmission of maternal microbiota and initial neonatal colonization, stool sampling as close to full term as possible, or at least within the third trimester of pregnancy, would have been ideal and most informative. This was not possible due to the need to incorporate biological sample collection into the established parent study. Vaginal microbiota analyses were not well powered. Our limited vaginal microbiota sample size (n = 28), which reduced study power, can be largely attributed to participant burden at the time of sampling as well as infant mortality, and exclusion of infants delivered via Caesarean section or without confirmed diagnoses from the FASD analyses. The use of 16S rRNA gene hypervariable sequencing limited taxonomic resolution to genus level, preventing species-and strain-level characterisation, and restricted functional inferences to *PICRUSt2* predictions, which should be interpreted with caution. Nevertheless, this study, conducted in a region with the highest documented FASD prevalence globally and historically accurate self-reported alcohol use (May et al., 2022, 2021), provides an important first step in informing and directing future hypothesis testing. We recommend that future well-powered studies utilize whole genome metagenomic sequencing to improve resolution at lower taxonomic levels and facilitate the direct measurement of microbial functional potential. The integration of metabolomic analysis, with specific focus on SCFAs and metabolites of the OCM, could serve to further functionally validate and extend the current findings.

## Conclusion

This is the first human study to evaluate maternal gut and vaginal bacterial composition in relation to prenatal alcohol exposure and infant FASD diagnosis. Our findings suggest that alcohol use during pregnancy disrupts the maternal gut microbiota, with potential implications for FASD development in exposed infants. While promising, validation in larger, well-powered longitudinal studies employing higher-resolution taxonomic and functional methodologies is needed. If replicable associations between the maternal microbiota, prenatal alcohol exposure, and infant FASD diagnosis are found, development of tractable therapeutic microbiota-targeted strategies could improve maternal and infant health.

## Declarations

### Ethics approval and consent to participate

This study was approved by the Stellenbosch University Health Research Ethics Committee (N13/01/013 and S19/07125). Written informed consent was obtained from all participants who were interviewed, assessed, and provided biological samples.

### Consent for publication

Not applicable

### Availability of data and materials

The data that support the findings of this study are available from the corresponding author upon reasonable request. The data are not publicly available due to privacy or ethical restrictions. Scripts supporting the analysis reported here are available at https://github.com/laurencmartin/maternal-microbiota-alcohol-fasd.

### Authors’ contributions

SMJH and SS conceptualized the study. A-SM supervised field staff and oversaw protocols and data collection. MMdV supervised data collection and aggregation in the field. NK was responsible for the selection, extraction, and preparation of the samples for sequencing. LCM, in collaboration with NK, KNVZ, MJD, and RK, performed the data analysis and statistical analyses included in the manuscript, LCM devised the design and content of the first draft of the manuscript. JSW and SMJH assisted with the conception, design and content of the manuscript and critical revision of the final draft. SS, PAM, LJH, JSW and SMJH provided senior guidance of the study as a whole and assisted with critical revision of the final draft. Each author contributed to, read, edited, and approved the contents of this manuscript.

## Data Availability

All data produced in the present study are available upon reasonable request to the authors.

## Acknowledgements

We extend our sincere gratitude to the participants who generously provided biological samples and shared sensitive health information during their pregnancies. We are also grateful to the Fetal Alcohol Syndrome Epidemiology Research (FASER) team, headed by Anna-Susan Marais and Marlene de Vries, for assistance in sample collection and their invaluable insight.

We acknowledge the use of the ilifu cloud computing facility, a partnership between the University of Cape Town, the University of the Western Cape, Stellenbosch University, Sol Plaatje University, and the Cape Peninsula University of Technology. The ilifu facility is supported by contributions from the Inter-University Institute for Data Intensive Astronomy (IDIA - a partnership between the University of Cape Town, the University of Pretoria, and the University of the Western Cape), the Computational Biology division at UCT, and the Data Intensive Research Initiative of South Africa (DIRISA).

