## Supporting Information for "Maternal gut and vaginal microbiota, alcohol use during pregnancy, and infant fetal alcohol spectrum disorder diagnosis in a South African cohort"

#### Supporting Tables, Data & Figures

Lauren C. Martin[1,2, *], Natasha Kitchin[1,2], Jacqueline S. Womersley[1,2,3,4], Kristien Nel Van Zyl[5], Anna-Susan Marais[1], Marlene M. de Vries[1], Matthew J. Dalby[6,7], Raymond Kiu[6,7,8], Lindsay J. Hall[6,7,8,9], Philip A. May[10], Soraya Seedat[1,2], Sian M.J. Hemmings[1,2, *]

[1] Department of Psychiatry, Faculty of Medicine and Health Sciences, Stellenbosch University, Cape Town, South Africa

[2] South African Medical Research Council/Stellenbosch University Genomics of Brain Disorders Research Unit, Stellenbosch University, Cape Town, South Africa

[3] Neuroscience Institute, University of Cape Town, Cape Town, South Africa

[4] Department of Human Biology, Faculty of Health Sciences, University of Cape Town, Cape Town, South Africa

[5] African Microbiome Institute, Division of Molecular Biology and Human Genetics, Department of Biomedical Sciences, Faculty of Medicine and Health Sciences, Stellenbosch University, Cape Town, South Africa

[6] Institute of Microbiology and Infection, University of Birmingham, Birmingham, United Kingdom

[7] Department of Microbes, Infection and Microbiomes, School of Infection, Inflammation and Immunology, College of Medicine and Health, University of Birmingham, Birmingham, United Kingdom

[8] Food, Microbiome and Health, Quadram Institute Bioscience, Norwich Research Park, Norwich, United Kingdom

[9] Norwich Medical School, University of East Anglia, Norwich Research Park, Norwich, United Kingdom

[10] Department of Nutrition, Gillings School of Global Public Health, Nutrition Research Institute, University of North Carolina, United States of America

Martin; Kitchin; Womersley; Nel Van Zyl; Marais; de Vries; Dalby; Kiu; Hall; May; Seedat; Hemmings

Corresponding authors: Lauren C. Martin & Sian M.J. Hemmings*

Correspondence address: Department of Psychiatry, Faculty of Medicine and Health Sciences, Stellenbosch University, PO Box 241, Cape Town 8000, South Africa

**Table S1:** Covariate adjustment sets for maternal fecal and vaginal microbiota alcohol use and infant FASD diagnosis analyses.

| **Diversity metric** | **Covariate** | **Effect size** | ***p*-value** |
|---|---|---|---|
| **Alcohol use maternal fecal microbiota analysis (n = 207)** | | | |
| Bray-Curtis dissimilarity | GA | 0.0164 | 0.001 |
| Bray-Curtis dissimilarity | BSS | 0.0210 | 0.003 |
| Bray-Curtis dissimilarity | Educational attainment | 0.0170 | 0.008 |
| Bray-Curtis dissimilarity | Extraction batch | 0.0673 | 0.023 |
| Observed richness | BSS | 3.4500 | 0.034 |
| Bray-Curtis dissimilarity | BMI | 0.0107 | 0.042 |
| Observed richness | Smoking | 3.9000 | 0.050 |
| ~ **AUDIT/HAU** + GA + BSS + Educational attainment + Extraction batch + BMI + Smoking | | | |
| **Infant FASD diagnosis maternal fecal microbiota analysis (n = 168)** | | | |
| Bray-Curtis dissimilarity | GA | 0.0183 | 0.007 |
| Bray-Curtis dissimilarity | Educational attainment | 0.0166 | 0.012 |
| Bray-Curtis dissimilarity | Extraction batch | 0.0854 | 0.014 |
| Bray-Curtis dissimilarity | BSS | 0.0257 | 0.020 |
| Bray-Curtis dissimilarity | Antibiotics | 0.0131 | 0.048 |
| Shannon diversity | BSS | 3.5100 | 0.032 |
| Observed richness | HIV | 4.3600 | 0.038 |
| Observed richness | BSS | 3.4700 | 0.033 |
| Bray-Curtis dissimilarity | BMI | 0.0139 | 0.040 |
| ~ **Infant diagnosis group** + GA + Educational attainment + Extraction batch + Antibiotics + BSS + HIV + BMI | | | |
| **Alcohol use maternal vaginal microbiota analysis (n = 28)** | | | |
| Observed richness | Age | 1.5000 | 0.009 |
| **~ AUDIT/HAU + Age** | | | |
| **Infant FASD diagnosis maternal vaginal microbiota analysis (n = 15)** | | | |
| Observed richness | Smoking | 9.6100 | 0.008 |
| Observed richness | Age* | -1.1590 | 0.050 |
| **~ Infant diagnosis group + Smoking** | | | |

*Variable not included due to constrained sample size and the risk of statistical overfitting.
AUDIT – Alcohol Use Disorders Identification Test; BMI – body mass index; BSS – Bristol Stool Scale; GA – Gestational age; HAU – hazardous alcohol use; HIV - Human Immunodeficiency Virus

**Table S2:** Characteristics of participants in the cross-sectional maternal fecal and vaginal microbiota infant FASD diagnosis investigation.

| | **Participants that provided fecal samples** | | | | | **Participants that provided vaginal swab samples** | | | | |
|---|---|---|---|---|---|---|---|---|---|---|
| | **All participants (n = 168)** | **FASD[a] (n = 80)** | **Not FASD (n = 88)** | **t/W/$X^2$ value** | ***p*-value** | **All participants (n = 15)** | **FASD[a] (n = 10)** | **Not FASD (n = 5)** | **t/W/$X^2$ value** | ***p*-value** |
| **Clinical and socio-demographic information** | | | | | | | | | | |
| **Age (years)** | 26.00 (22.00 – 27.21) | 25.50 (22.00 – 33.00) | 26.00 (22.00 – 31.00) | 3651 | 0.678 | 29.47 (5.33) | 30.10 (5.36) | 28.20 (5.63) | 0.63 | 0.549 |
| **BMI (kg/$m^2$)** | 24.90 (21.07 – 30.98) | 23.65 (21.00 – 28.95) | 25.85 (21.20 – 31.78) | 3085 | 0.168 | 22.67 (2.98) | 22.30 (3.19) | 23.41 (2.68) | -0.71 | 0.496 |
| **Gestational age at sample collection (weeks)** | 19.00 (14.00 – 26.00) | 20.00 (14.00 – 26.25) | 18.00 (13.50 – 24.00) | 3873.50 | 0.262 | 38.13 (1.41) | 38.30 (1.25) | 37.80 (1.79) | 0.56 | 0.596 |
| **Antibiotic use in past six months, *n (%)*** | 25 (14.88) | 10 (12.50) | 15 (17.05) | 0.37 | 0.542 | 2 (13.33) | 2 (20.00) | 0.00 | - | 0.524 |
| **HIV-positive serostatus, *n (%)*** | 6 (3.57) | 3 (3.75) | 3 (3.41) | 1.07e-29 | 1.000 | 2 (13.33) | 2 (20.00) | 0.00 | - | 0.524 |
| **Maternal education (highest grade completed)** | 9.00 (7.00 – 11.00) | 8.00 (7.00 – 10.00) | 9.00 (8.00 – 11.00) | 2700.50 | **0.009** | 8.60 (2.26) | 8.10 (2.51) | 9.60 (1.34) | -1.51 | 0.156 |
| **Employment status, *n (% employed)*** | 67 (39.88) | 30 (37.50) | 37 (42.05) | 0.20 | 0.658 | 5 (33.33) | 4 (40.00) | 1 (20.00) | - | 0.600 |
| **Alcohol and substance use during index pregnancy** | | | | | | | | | | |
| **AUDIT score** | 11.00 (0.00 – 17.00) | 15.00 (8.00 – 21.00) | 4.50 (0.00 – 12.25) | 5074.50 | **< 0.001** | 17.27 (10.68) | 20.80 (10.49) | 10.20 (7.63) | -2.23 | **0.048** |
| **HAU, *n (%)*[b]** | 108 (64.29) | 68 (85.00) | 40 (45.45) | 26.85 | **< 0.001** | 13 (86.67) | 10 (100.00) | 3 (60.00) | - | 0.600 |
| **PEth level (ng/mL)[c,d]** | 15.00 (0.00 – 75.33) | 44.84 (5.75 – 210.50) | 27.11 (5.00 – 23.84) | 3955 | **< 0.001** | 107.00 (79.00 – 153.50) | 107.00 (79.00 – 153.50) | **-** | **-** | **-** |
| **EtG level (pg/mg)[e,f]** | 0.00 (0.00 – 30.00) | 19.00 (0.00 – 76.50) | 0.00 (0.00 – 6.00) | 3467 | **< 0.001** | 0.00 (0.00 – 36.25) | 0.00 (0.00 – 36.25) | **-** | **-** | **-** |
| **Smoked during index pregnancy, *n (%)*** | 111 (66.07) | 59 (73.75) | 52 (59.09) | 3.39 | 0.066 | 12 (80.00) | 8 (80.00) | 4 (80.00) | - | 1.000 |
| **Characteristics of child born from index pregnancy** | | | | | | | | | | |
| **Sex, *n (% male)*** | 86 (51.19) | 45 (56.25) | 41 (46.59) | 0.00 | 1.000 | 7 (46.67) | 4 (40.00) | 3 (30.00) | - | 0.608 |

Continuous data are summarised as mean (standard deviation) if normally distributed, and as median (interquartile range) if non-normally distributed, unless otherwise specified. Categorical data are summarised as counts and percentages.
[a] Participants were included in the FASD group on the basis of their infant receiving a confirmed diagnosis on the FASD spectrum.
[b] Participants were considered to have consumed alcohol hazardously during pregnancy if AUDIT score ≥ 7.
[c] PEth levels were available for 72 participants in the FASD group and 79 participants in the Not FASD group for the fecal analysis.
[d] PEth levels were available for 5 participants in the FASD group and 0 participants in the Not FASD group for the vaginal analysis.
[e] EtG levels were available for 69 participants in the FASD group and 71 participants in the Not FASD group for the fecal analysis.
[f] EtG levels were available for 5 participants in the FASD group 0 participants in the Not FASD group for the vaginal analysis.
AUDIT - Alcohol Use Disorders Identification Test; BMI - body mass index; EtG – Ethyl Glucuronide; FASD – fetal alcohol spectrum disorders; HAU – hazardous alcohol consumption; HIV - Human Immunodeficiency Virus; PEth – Phosphatidylethanol

**Table S3:** Phylum-level median relative bacterial abundance of maternal fecal samples in alcohol use (n = 207) and infant FASD diagnosis analysis (n = 168).

| **Alcohol use fecal analysis (n = 207)** | | **FASD diagnosis fecal analysis (n = 168)** | |
|---|---|---|---|
| **Phylum** | **Median relative abundance (%)** | **Phylum** | **Median relative abundance (%)** |
| Bacillota | 45.10 | Bacillota | 45.40 |
| Bacteroidota | 38.50 | Bacteroidota | 39.00 |
| Pseudomonadota | 6.96 | Pseudomonadota | 6.49 |
| Actinomycetota | 1.65 | Actinomycetota | 1.62 |
| Thermodesulfobacteriota | 0.57 | Thermodesulfobacteriota | 0.54 |
| Verrucomicrobiota | 0.01 | Verrucomicrobiota | 0.009 |
| Cyanobacteriota | 0.008 | Cyanobacteriota | 0.007 |

**Table S4:** Median relative bacterial abundance of top ten genera in maternal fecal samples in alcohol use (n = 207) and infant FASD diagnosis analysis (n = 168).

| **Alcohol use fecal analysis (n = 207)** | | **FASD diagnosis fecal analysis (n = 168)** | |
|---|---|---|---|
| **Genus** | **Median relative abundance (%)** | **Genus** | **Median relative abundance (%)** |
| *Segatella* | 23.30 | *Segatella* | 21.30 |
| *Faecalibacterium* | 4.42 | *Faecalibacterium* | 4.55 |
| *Succinivibrio* | 3.86 | *Oscillospiraceae UCG-002* | 3.70 |
| *Oscillospiraceae UCG-002* | 3.61 | *Succinivibrio* | 3.56 |
| *Bacteroides* | 3.09 | *Bacteroides* | 3.32 |
| *Dialister* | 1.90 | *Dialister* | 1.57 |
| *Leyella* | 1.51 | *Leyella* | 1.21 |
| *Lachnospiraceae NK4A136* group | 1.16 | *Lachnospiraceae NK4A136* group | 1.23 |
| *Roseburia* | 1.08 | *Roseburia* | 1.07 |
| *Sutterella* | 1.06 | *Oscillospiraceae UCG-005* | 1.06 |

The most abundant phyla across all the fecal samples included in the FASD diagnosis analysis (n = 168) were *Bacillota* (formerly *Firmicutes*), *Bacteroidota* (formerly *Bacteroidetes*), *Pseudomonadota*, *Actinomycetota*, *Thermodesulfobacteriota*, *Verrucomicrobiota* and *Cyanobacteriota* (**Table S2**).

At the genus-level, the fecal microbiota of the 168 participants who provided fecal samples and had an infant who had undergone diagnostic assessment for FASD (Aim 2) was dominated by *Segatella* species, which was present at a median relative abundance of 21.3% (**Table S3**). In addition, and listed in order of decreasing median abundance, *Faecalibacterium* (4.6%), *Oscillospiraceae UCG-002* (3.7%), *Succinivibrio* (3.6%), *Bacteroides* (3.3%), *Dialister* (1.6%), *Lachnospiraceae NK4A136* group (1.2%), *Leyella* (1.2%), *Roseburia* (1.1%) and *Oscillospiraceae UCG-005* (1.1%), were among the remaining top ten most abundant bacterial genera present.

**Table S5:** Maternal fecal genera with the strongest nominal joint associations (*p*-joint) with self-reported alcohol use measures (AUDIT score; AUDIT ≥ 7) and infant FASD diagnostic group, identified using MaAsLin3's linear mixed-effects abundance model and logistic prevalence model.

| | Abundance model | | | | | | | Prevalence model | | | | | | |
|---|---|---|---|---|---|---|---|---|---|---|---|---|---|---|
| **Genus** | **Coef[a]** | **SE** | ***p*-abun** | ***q*-abun** | **N[*]** | ***p*-joint** | ***q*-joint** | **Coef[b]** | **SE** | ***p*-abun** | ***q*-abun** | **N[*]** | ***p*-joint** | ***q*-joint** |
| **Continuous AUDIT score (n = 207)** | | | | | | | | | | | | | | |
| *Victivallis* | -0.408 | 0.172 | **0.017** | 0.402 | 111 | 0.002 | 0.319 | -0.566 | 0.168 | **0.001** | 0.306 | 111 | 0.002 | 0.319 |
| *Butyricimonas* | 0.011 | 0.160 | 0.986 | 0.999 | 135 | 0.004 | 0.319 | -0.535 | 0.172 | **0.002** | 0.306 | 135 | 0.004 | 0.319 |
| *Erysipelatoclostridiaceae UCG-004* | 0.096 | 0.211 | 0.696 | 0.946 | 115 | 0.006 | 0.319 | -0.515 | 0.174 | **0.003** | 0.306 | 115 | 0.006 | 0.319 |
| *Erysipelotrichaceae UCG-003* | 0.196 | 0.299 | 0.542 | 0.918 | 77 | 0.007 | 0.319 | 0.490 | 0.168 | **0.004** | 0.306 | 77 | 0.007 | 0.319 |
| *Oscillospiraceae NK4A214* group | -0.339 | 0.119 | **0.004** | 0.313 | 198 | 0.009 | 0.326 | -0.080 | 0.360 | 0.825 | 0.976 | 198 | 0.009 | 0.326 |
| *Oscillospiraceae UCG-005* | -0.466 | 0.166 | **0.005** | 0.313 | 199 | 0.010 | 0.326 | -0.184 | 0.379 | 0.627 | 0.943 | 199 | 0.010 | 0.326 |
| *Oscillospiraceae UCG-002* | -0.388 | 0.142 | **0.006** | 0.334 | 199 | 0.012 | 0.341 | -0.058 | 0.409 | 0.887 | 0.986 | 199 | 0.012 | 0.341 |
| *Butyricicoccaceae UCG-009* | -0.165 | 0.203 | 0.382 | 0.832 | 130 | 0.017 | 0.341 | -0.454 | 0.173 | 0.009 | 0.334 | 130 | 0.017 | 0.341 |
| *Lachnospiraceae [Eubacterium] oxidoreducens* group | 0.492 | 0.176 | **0.010** | 0.334 | 66 | 0.019 | 0.341 | -0.371 | 0.174 | **0.033** | 0.450 | 66 | 0.019 | 0.341 |
| *Mediterraneibacter* | 0.368 | 0.135 | **0.011** | 0.334 | 183 | 0.022 | 0.341 | -0.340 | 0.258 | 0.188 | 0.686 | 183 | 0.022 | 0.341 |
| *Intestinibacter* | -0.432 | 0.181 | **0.016** | 0.402 | 103 | 0.033 | 0.408 | 0.036 | 0.161 | 0.823 | 0.976 | 103 | 0.033 | 0.408 |
| *Bacteroides* | 0.231 | 0.232 | 0.351 | 0.813 | 202 | 0.035 | 0.411 | -1.933 | 0.816 | **0.018** | 0.402 | 202 | 0.035 | 0.411 |
| *Lachnospiraceae [Eubacterium] xylanophilum* group | 0.029 | 0.208 | 0.942 | 0.991 | 109 | 0.040 | 0.411 | -0.380 | 0.164 | **0.020** | 0.402 | 109 | 0.040 | 0.411 |
| *Ruminococcus* | 0.040 | 0.131 | 0.841 | 0.979 | 199 | 0.047 | 0.426 | -1.212 | 0.536 | **0.024** | 0.421 | 199 | 0.047 | 0.426 |
| *Desulfovibrio* | -0.171 | 0.124 | 0.144 | 0.649 | 171 | 0.048 | 0.430 | -0.473 | 0.210 | **0.025** | 0.421 | 171 | 0.048 | 0.430 |
| **HAU[c] ( n = 207)** | | | | | | | | | | | | | | |
| *Victivallis* | -0.540 | 0.341 | 0.098 | 0.641 | 111 | 0.008 | 0.359 | -1.016 | 0.354 | **0.004** | 0.346 | 111 | 0.008 | 0.359 |
| *Erysipelotrichaceae UCG-003* | 0.173 | 0.660 | 0.834 | 0.987 | 77 | 0.014 | 0.359 | 1.063 | 0.393 | **0.007** | 0.346 | 77 | 0.014 | 0.359 |
| *Lachnospiraceae [Eubacterium] oxidoreducens* group | 0.962 | 0.370 | **0.016** | 0.430 | 66 | 0.032 | 0.420 | -0.394 | 0.357 | 0.269 | 0.765 | 66 | 0.032 | 0.420 |
| *Intestinibacter* | -0.918 | 0.388 | **0.017** | 0.430 | 103 | 0.034 | 0.420 | 0.323 | 0.345 | 0.350 | 0.821 | 103 | 0.034 | 0.420 |
| *Paraprevotella* | 0.827 | 0.328 | **0.019** | 0.430 | 99 | 0.038 | 0.420 | -0.397 | 0.346 | 0.251 | 0.765 | 99 | 0.038 | 0.420 |
| *Roseburia* | 0.635 | 0.253 | **0.021** | 0.443 | 197 | 0.042 | 0.435 | -1.018 | 0.865 | 0.239 | 0.753 | 197 | 0.042 | 0.435 |
| *Clostridium* | 0.147 | 0.353 | 0.751 | 0.977 | 179 | 0.043 | 0.435 | -1.415 | 0.617 | 0.022 | 0.443 | 179 | 0.043 | 0.435 |
| **Infant FASD diagnostic category (n = 168)** | | | | | | | | | | | | | | |
| *Lachnospiraceae CAG-56* | -0.415 | 0.362 | 0.423 | 0.872 | 73 | 0.009 | 0.458 | -0.962 | 0.339 | **0.005** | 0.438 | 73 | 0.009 | 0.458 |
| *Lachnospiraceae [Eubacterium] xylano philum* group | -0.078 | 0.383 | 0.905 | 0.994 | 94 | 0.009 | 0.458 | -0.957 | 0.339 | **0.005** | 0.438 | 94 | 0.009 | 0.458 |
| *Asteroleplasma*[#] | - | - | - | - | 48 | 0.010 | 0.458 | -0.956 | 0.372 | **0.010** | 0.438 | 48 | 0.010 | 0.458 |
| *Bifidobacterium* | -0.913 | 0.367 | **0.035** | 0.558 | 143 | 0.010 | 0.458 | -1.540 | 0.550 | **0.005** | 0.438 | 143 | 0.010 | 0.458 |
| *Lachnospiraceae ND3007* group | -0.741 | 0.255 | **0.019** | 0.508 | 140 | 0.014 | 0.458 | -1.242 | 0.459 | **0.007** | 0.438 | 140 | 0.014 | 0.458 |
| *Butyribacter* | -0.627 | 0.455 | 0.274 | 0.802 | 79 | 0.017 | 0.458 | -0.952 | 0.362 | **0.008** | 0.438 | 79 | 0.017 | 0.458 |
| *Negativibacillus* | 0.794 | 0.340 | **0.009** | 0.438 | 85 | 0.018 | 0.458 | -0.557 | 0.321 | 0.083 | 0.623 | 85 | 0.018 | 0.458 |

| | | | | | | | | | | | | | | |
|---|---|---|---|---|---|---|---|---|---|---|---|---|---|---|
| *Ruminococcaceae [Eubacterium] siraeum* group | 0.186 | 0.378 | 0.414 | 0.868 | 123 | 0.019 | 0.458 | -1.042 | 0.403 | **0.010** | 0.438 | 123 | 0.019 | 0.458 |
| *Coriobacteriaceae UCG-003*[#] | - | - | - | - | 26 | 0.021 | 0.458 | -1.161 | 0.501 | **0.021** | 0.508 | 26 | 0.021 | 0.458 |
| *Fusicatenibacter* | -0.806 | 0.271 | **0.015** | 0.496 | 138 | 0.029 | 0.519 | -0.631 | 0.429 | 0.142 | 0.682 | 138 | 0.029 | 0.519 |
| *Catenibacterium* | -0.804 | 0.278 | **0.018** | 0.508 | 124 | 0.036 | 0.551 | 0.224 | 0.374 | 0.549 | 0.932 | 124 | 0.036 | 0.551 |
| *Christensenellaceae R-7* group | -0.060 | 0.401 | 0.873 | 0.994 | 157 | 0.042 | 0.551 | -2.698 | 1.173 | **0.021** | 0.508 | 157 | 0.042 | 0.551 |

Co-efficients (Coef) represent the difference in each group relative to the Control group for the categorical measures.
[a] Abundance co-efficient representing the log2 fold per one-unit change in continuous measure or between-group difference for a given feature, if present.
[b] Prevalence co-efficient representing the log-odds ratio of a feature's presence between groups or for every one-unit increase in continuous measure.
[c] Participants were considered to have consumed alcohol hazardously during pregnancy if AUDIT score ≥ 7.
* Number of samples that did not have a zero value.
[#] MaAsLin3 linear abundance model could not be fitted.
AUDIT – Alcohol Use Disorders Identification Test; FASD – fetal alcohol spectrum disorder; HAU – hazardous alcohol use; SE - standard error; q-value – Benjamini-Hochberg false discovery rate (FDR)-adjusted p-value

**Table S6:** *PICRUSt2*-predicted maternal fecal microbiota KEGG pathways nominally associated with alcohol use and infant FASD diagnosis

| Outcome | KEGG pathway | Category | Sub-category | Pathway description | Coef (β)* | SE | *p*-value | *q*-value |
|---|---|---|---|---|---|---|---|---|
| **AUDIT score (n = 207)** | ko00552 | Metabolism | Glycan biosynthesis and metabolism | Teichoic acid biosynthesis | -0.030 | 0.009 | 0.007 | 0.363 |
| | ko00040 | Metabolism | Carbohydrate metabolism | Pentose and glucuronate interconversions | 0.033 | 0.015 | 0.017 | 0.479 |
| | ko99987 | - | - | - | 0.102 | 0.046 | 0.026 | 0.532 |
| | ko00785 | Metabolism | Metabolism of cofactors and vitamins | Lipoic acid metabolism | -0.133 | 0.058 | 0.027 | 0.532 |
| | ko00591 | Metabolism | Lipid metabolism | Linoleic acid metabolism | -0.248 | 0.112 | 0.031 | 0.532 |
| | ko03051 | - | - | - | -0.020 | 0.007 | 0.032 | 0.532 |
| | ko04614 | Organismal systems | Endocrine system | Renin-angiotensin system | -0.173 | 0.079 | 0.033 | 0.532 |
| | ko03010 | Genetic information processing | Translation | Ribosome | -0.022 | 0.008 | 0.033 | 0.532 |
| | ko03011 | - | - | - | -0.022 | 0.008 | 0.033 | 0.532 |
| | ko99977 | - | - | - | 0.023 | 0.011 | 0.033 | 0.532 |
| | ko00565 | Metabolism | Lipid metabolism | Ether lipid metabolism | -0.230 | 0.106 | 0.033 | 0.532 |
| | ko00410 | Metabolism | Metabolism of other amino acids | beta-Alanine metabolism | 0.021 | 0.010 | 0.034 | 0.533 |
| | ko00970 | Genetic information processing | Translation | Aminoacyl-tRNA biosynthesis | -0.023 | 0.009 | 0.036 | 0.556 |
| | ko99994 | - | - | - | 0.0333 | 0.017 | 0.037 | 0.561 |
| | ko00670 | Metabolism | Metabolism of cofactors and vitamins | One carbon pool by folate | -0.023 | 0.009 | 0.038 | 0.561 |
| | ko00480 | Metabolism | Metabolism of other amino acids | Glutathione metabolism | 0.023 | 0.012 | 0.040 | 0.561 |
| | ko03420 | Genetic information processing | Replication and repair | Nucleotide excision repair | -0.021 | 0.008 | 0.041 | 0.561 |
| | ko00514 | Metabolism | Glycan biosynthesis and metabolism | Other types of O-glycan biosynthesis | -0.186 | 0.090 | 0.043 | 0.561 |
| | ko03016 | - | - | - | -0.020 | 0.008 | 0.045 | 0.561 |
| | ko00052 | Metabolism | Carbohydrate metabolism | Galactose metabolism | 0.016 | 0.009 | 0.045 | 0.561 |
| | ko03430 | Genetic information processing | Replication and repair | Mismatch repair | -0.022 | 0.009 | 0.046 | 0.561 |
| | ko99996 | - | - | - | 0.022 | 0.012 | 0.047 | 0.561 |
| | ko00515 | Metabolism | Glycan biosynthesis and metabolism | Mannose type O-glycan biosynthesis | -0.163 | 0.080 | 0.048 | 0.561 |
| **HAU# (n = 207)** | ko04614 | Organismal systems | Endocrine system | Renin-angiotensin system | -0.348 | 0.169 | 0.040 | 0.593 |
| | ko00440 | Metabolism | Metabolism of other amino acids | Phosphonate and phosphinate metabolism | 0.090 | 0.044 | 0.048 | 0.593 |
| **Infant FASD diagnostic group (n = 168)** | ko05150 | Human diseases | Infectious disease: bacterial | *Staphylococcus aureus* infection | -0.418 | 0.135 | 0.003 | 0.259 |
| | ko00906 | Metabolism | Metabolism of terpenoids and polyketides | Carotenoid biosynthesis | -0.342 | 0.138 | 0.016 | 0.523 |
| | ko00552 | Metabolism | Glycan biosynthesis and metabolism | Teichoic acid biosynthesis | -0.059 | 0.020 | 0.016 | 0.523 |
| | ko99985 | - | - | - | 0.083 | 0.036 | 0.019 | 0.553 |
| | ko00997 | Metabolism | Biosynthesis of other secondary metabolites | Biosynthesis of various other secondary metabolites | -0.553 | 0.240 | 0.024 | 0.553 |
| | ko00542 | Metabolism | Glycan biosynthesis and metabolism | O-Antigen repeat unit biosynthesis | -0.300 | 0.131 | 0.029 | 0.553 |
| | ko00572 | Metabolism | Glycan biosynthesis and metabolism | Arabinogalactan biosynthesis - *Mycobacterium* | -0.293 | 0.131 | 0.030 | 0.553 |
| | ko00515 | Metabolism | Glycan biosynthesis and metabolism | Mannose type O-glycan biosynthesis | -0.341 | 0.158 | 0.036 | 0.553 |
| | ko04138 | Cellular processes | Transport and catabolism | Autophagy - yeast | -0.233 | 0.109 | 0.040 | 0.558 |

Differential abundance testing performed using MaAsLin3 v.0.99.16 linear models with total sum scaling and log transformation.
Statistical significance defined as q ≤ 0.10. Results with p < 0.05 and q > 0.10 are reported as nominal associations.
* Coefficient (Coef or β) represents the difference in the log-transformed relative abundance of the reported KEGG pathway per one-unit increase in continuous measures or between groups.
### Participants were considered to have consumed alcohol hazardously during pregnancy if AUDIT score ≥ 7.
AUDIT – Alcohol Use Disorders Identification Test; FASD – fetal alcohol spectrum disorder; HAU – hazardous alcohol use; SE - standard error; q-value – Benjamini-Hochberg false discovery rate (FDR)-adjusted p-value

**Table S7:** Maternal fecal microbiota MetaCyc pathways nominally associated with maternal alcohol use measures and infant FASD diagnosis.

| Outcome | MetaCyc ID | Class | Sub-class | Pathway | Co-eff (β)* | SE | *p*-value | *q*-value |
|---|---|---|---|---|---|---|---|---|
| **AUDIT score (n = 207)** | LIPASYN-PWY | Degradation, Utilization, Assimilation | Fatty acid and lipid degradation | Phospholipases | -0.349 | 0.114 | 0.003 | 0.121 |
| | PWY-8187 | Generation of precursor metabolites and energy; Degradation, Utilization, Assimilation | Fermentation; amino acid degradation | L-arginine degradation XIII (reductive Stickland reaction) | -0.216 | 0.073 | 0.005 | 0.148 |
| | POLYAMINSYN3-PWY | Biosynthesis | Amide, Amidine, Amine, and Polyamine Biosynthesis | Superpathway of polyamine biosynthesis II | -0.178 | 0.063 | 0.008 | 0.179 |
| | PWY-6471 | Biosynthesis; Detoxification | Cell structure biosynthesis; Antibiotic resistance | Peptidoglycan biosynthesis IV (*Enterococcus faecium*) | -0.215 | 0.085 | 0.016 | 0.259 |
| | PWY-4984 | Degradation, Utilization, Assimilation | Inorganic nutrient metabolism | Urea cycle | -0.200 | 0.083 | 0.021 | 0.281 |
| | PWY-6803 | Biosynthesis | Fatty acid and lipid biosynthesis | Phosphatidylcholine acyl editing | -0.250 | 0.108 | 0.027 | 0.313 |
| | PWY-7282 | Biosynthesis | Co-factor, carrier & vitamin biosynthesis | 4-amino-2-methyl-5-diphosphomethylpyrimidine biosynthesis II | 0.253 | 0.123 | 0.036 | 0.347 |
| | PWY-5265 | Biosynthesis | Cell structure biosynthesis | Peptidoglycan biosynthesis II (*staphylococci*) | -0.218 | 0.100 | 0.036 | 0.347 |
| | PWY0-845 | Biosynthesis | Co-factor, carrier & vitamin biosynthesis | Superpathway of pyridoxal 5'-phosphate biosynthesis and salvage | 0.265 | 0.134 | 0.044 | 0.367 |
| | PWY-7663 | Biosynthesis | Fatty acid and lipid biosynthesis | Gondoate biosynthesis (anaerobic) | 0.014 | 0.009 | 0.045 | 0.367 |
| **HAU# (n = 207)** | PWY-7820 | Biosynthesis | Cell structure biosynthesis | Teichuronic acid biosynthesis (*B. subtilis* 168) | -1.594 | 0.453 | **< 0.001** | **0.043** |
| | FUC-RHAMCAT-PWY | - | - | Superpathway of fucose and rhamnose degradation | 0.213 | 0.095 | 0.023 | 0.273 |
| **Infant FASD diagnostic group (n = 168)** | PWY-6470 | Biosynthesis; Detoxification | Cell-Structure-Biosynthesis; Antibiotic-Resistance | Peptidoglycan biosynthesis V (β-lactam resistance) | -0.335 | 0.100 | 0.002 | 0.104 |
| | PWY-7399 | Degradation, Utilization, Assimilation | Inorganic nutrient metabolism | Methylphosphonate degradation II | -0.953 | 0.306 | 0.003 | 0.110 |
| | PWY-6383 | Biosynthesis | Co-factor, carrier & vitamin biosynthesis; Polyprenyl biosynthesis | Mono-trans, poly-cis decaprenyl phosphate biosynthesis | -0.669 | 0.253 | 0.012 | 0.265 |
| | HOMOSER-METSYN-PWY | Biosynthesis | Amino acid biosynthesis | L-methionine biosynthesis I | -0.298 | 0.111 | 0.015 | 0.297 |
| | PWY-5347 | Biosynthesis | Amino acid biosynthesis | Superpathway of L-methionine biosynthesis (transulfuration) | -0.272 | 0.100 | 0.015 | 0.297 |
| | METSYN-PWY | Biosynthesis | Amino acid biosynthesis | Superpathway of L-homoserine and L-methionine biosynthesis | -0.277 | 0.102 | 0.015 | 0.297 |
| | GLYCOLYSIS-E-D | Generation of precursor metabolites and energy | - | Superpathway of glycolysis and the Entner-Doudoroff pathway | 0.822 | 0.345 | 0.016 | 0.298 |

| | | | | | | | |
|---|---|---|---|---|---|---|---|
| MET-SAM-PWY | Biosynthesis | Co-factor, carrier & vitamin biosynthesis | Superpathway of S-adenosyl-L-methionine biosynthesis | -0.260 | 0.098 | 0.018 | 0.300 |
| METH-ACETATE-PWY | Generation of precursor metabolites and energy; Degradation, Utilization, Assimilation | Respiration; Acetate degradation | Methanogenesis from acetate | -0.430 | 0.174 | 0.021 | 0.309 |
| PWY-8004 | Generation of precursor metabolites and energy; Degradation, Utilization, Assimilation | Entner-Doudoroff pathways; Carbohydrate degradation | Entner-Doudoroff pathway I | 0.762 | 0.336 | 0.021 | 0.309 |
| PWY-6292 | Biosynthesis | Amino acid biosynthesis | Superpathway of L-cysteine biosynthesis (mammalian) | -0.400 | 0.162 | 0.021 | 0.309 |
| PWY66-399 | Biosynthesis | Carbohydrate synthesis | Gluconeogenesis III | -0.416 | 0.173 | 0.024 | 0.328 |
| PPGPPMET-PWY | Biosynthesis | Metabolic regulator biosynthesis | ppGpp metabolism | 0.097 | 0.051 | 0.026 | 0.328 |
| PWY-6471 | Biosynthesis; Detoxification | Cell structure biosynthesis; Antibiotic resistance | Peptidoglycan biosynthesis IV (*Enterococcus faecium*) | -0.420 | 0.180 | 0.029 | 0.337 |
| ANAEROFRUCAT-PWY | Generation of precursor metabolites and energy | Fermentation | Homolactic fermentation | -0.234 | 0.097 | 0.032 | 0.338 |
| PWY-5100 | Generation of precursor metabolites and energy | Fermentation | Pyruvate fermentation to acetate and lactate II | -0.175 | 0.070 | 0.033 | 0.338 |
| KDO-NAGLIPASYN-PWY | Biosynthesis | Fatty acid & lipid biosynthesis; Cell structure biosynthesis | Superpathway of (Kdo)2-lipid A biosynthesis | 0.612 | 0.298 | 0.035 | 0.340 |
| PWY-6123 | Biosynthesis | Nucleoside & nucleotide biosynthesis | Inosine-5'-phosphate biosynthesis I | 0.027 | 0.018 | 0.037 | 0.340 |
| CODH-PWY | Degradation, Utilization, Assimilation | C1 compound utilization and assimilation | Reductive acetyl coenzyme A pathway I (homoacetogenic bacteria) | -0.385 | 0.172 | 0.037 | 0.341 |
| PWY-8187 | Generation of precursor metabolites and energy; Degradation, Utilization, Assimilation | Fermentation; Amino acid degradation | L-arginine degradation XIII (reductive Stickland reaction) | -0.330 | 0.149 | 0.041 | 0.365 |
| PWY-6121 | Biosynthesis | Nucleoside & nucleotide biosynthesis | 5-aminoimidazole ribonucleotide biosynthesis I | 0.022 | 0.017 | 0.049 | 0.393 |
| PWY-6124 | Biosynthesis | Nucleoside & nucleotide biosynthesis | Inosine-5'-phosphate biosynthesis II | 0.025 | 0.019 | 0.049 | 0.393 |

Differential abundance testing performed using MaAsLin3 v.0.99.16 linear models with total sum scaling and log transformation.
Statistical significance defined as $q \leq 0.10$ (text emboldened). Results with $p < 0.05$ and $q > 0.10$ are reported as nominal associations.
* Coefficient (Coef or β) represents the difference in the log-transformed relative abundance of each MetaCyc pathway per unit increase in continuous measures or between groups.
[#] Participants were considered to have consumed alcohol hazardously during pregnancy if AUDIT score ≥ 7.
AUDIT – Alcohol Use Disorders Identification Test; FASD – fetal alcohol spectrum disorder; HAU – hazardous alcohol use; SE - standard error; q-value – Benjamini-Hochberg false discovery rate (FDR)-adjusted *p*-value.

**Table S8:** Phylum-level median relative bacterial abundance of maternal vaginal swab samples in alcohol use (n = 28) and infant FASD diagnosis analysis (n = 15).

| **Alcohol use vaginal swab analysis (n = 28)** | | **FASD diagnosis vaginal swab analysis (n = 15)** | |
|---|---|---|---|
| **Phylum** | **Median relative abundance (%)** | **Phylum** | **Median relative abundance (%)** |
| *Bacillota* | 86.90 | *Bacillota* | 58.20 |
| *Actinomycetota* | 10.30 | *Actinomycetota* | 15.20 |
| *Bacteroidota* | 1.96 | *Bacteroidota* | 1.23 |
| *Fusobacteriota* | 0.02 | *Pseudomonadota* | 0.001 |
| *Pseudomonadota* | 0.01 | *Fusobacteriota* | 0.001 |

**Table S9:** Median relative bacterial abundance of top five genera in maternal vaginal swab samples in alcohol use (n = 28) and infant FASD diagnosis analysis (n = 15).

| **Alcohol use vaginal swab analysis (n = 28)** | | **FASD diagnosis vaginal swab analysis (n = 15)** | |
|---|---|---|---|
| **Genus** | **Median relative abundance (%)** | **Genus** | **Median relative abundance (%)** |
| *Lactobacillus* | 72.50 | *Lactobacillus* | 45.80 |
| *Gardnerella* | 5.28 | *Gardnerella* | 6.24 |
| *Prevotella* | 0.56 | *Prevotella* | 0.03 |
| *Ureaplasma* | 0.27 | *Fannyhessea* | 0.01 |
| *Hoylesella* | 0.21 | *Ureaplasma* | 0.01 |

The most abundant bacterial phyla present across the maternal vaginal swab samples, in order of decreasing abundance, were: *Bacillota*, *Actinomycetota*, *Bacteroidota*, *Fusobacteriota* and *Pseudomonadota* (**Table S8**).

At the genus-level, the vaginal microbiota of the 15 participants was dominated by *Lactobacillus* species, which was present at a median relative abundance of 45.8% (**Table S9**). In addition, and listed in order of decreasing median abundance, *Gardnerella* (6.2%), *Prevotella* (0.03%), *Fannyhessea* (0.01%), and *Ureaplasma* (0.01%) were among the remaining top five most abundant bacterial genera present in the samples.

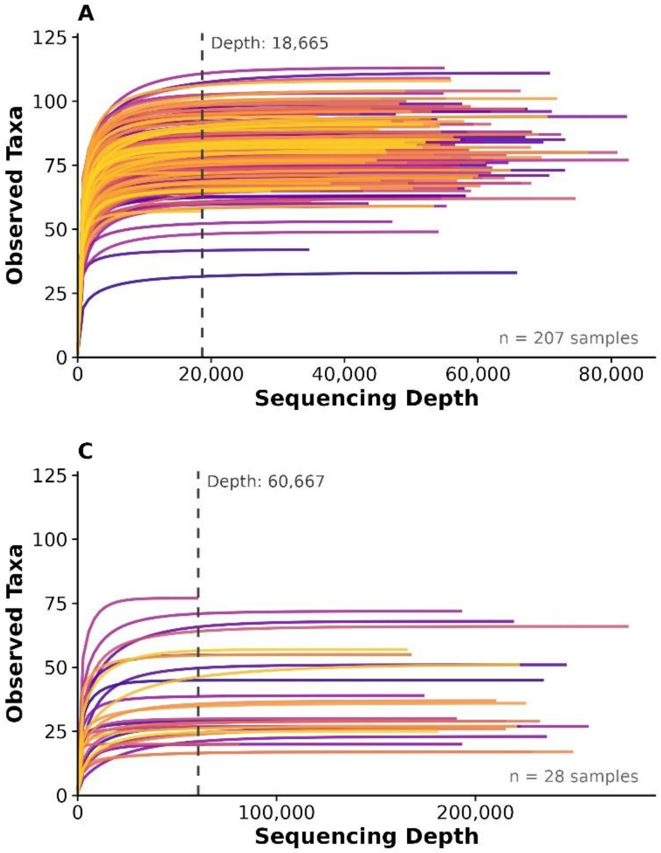


**Figure S1: Sequencing depth rarefaction curves for the maternal fecal and vaginal microbiota datasets across alcohol use and infant FASD diagnosis analyses.** Genus-level rarefaction curves are depicted for (**A**) fecal microbiota alcohol use analyses, (**B**) fecal microbiota infant FASD diagnosis analysis, (**C**) vaginal microbiota alcohol use analyses and (**D**) vaginal microbiota infant FASD diagnosis group analysis, conducted on fecal (n = 207) and vaginal swab (n = 28) samples collected from pregnant participants. The vertical line represents the rarefaction depth at which each dataset was sub-sampled. Reads were normalised by random sub-sampling without replacement to an even depth of 18,665 reads, 27,331 reads, 60,667 reads and 81,236 reads for the aforementioned analyses, respectively.

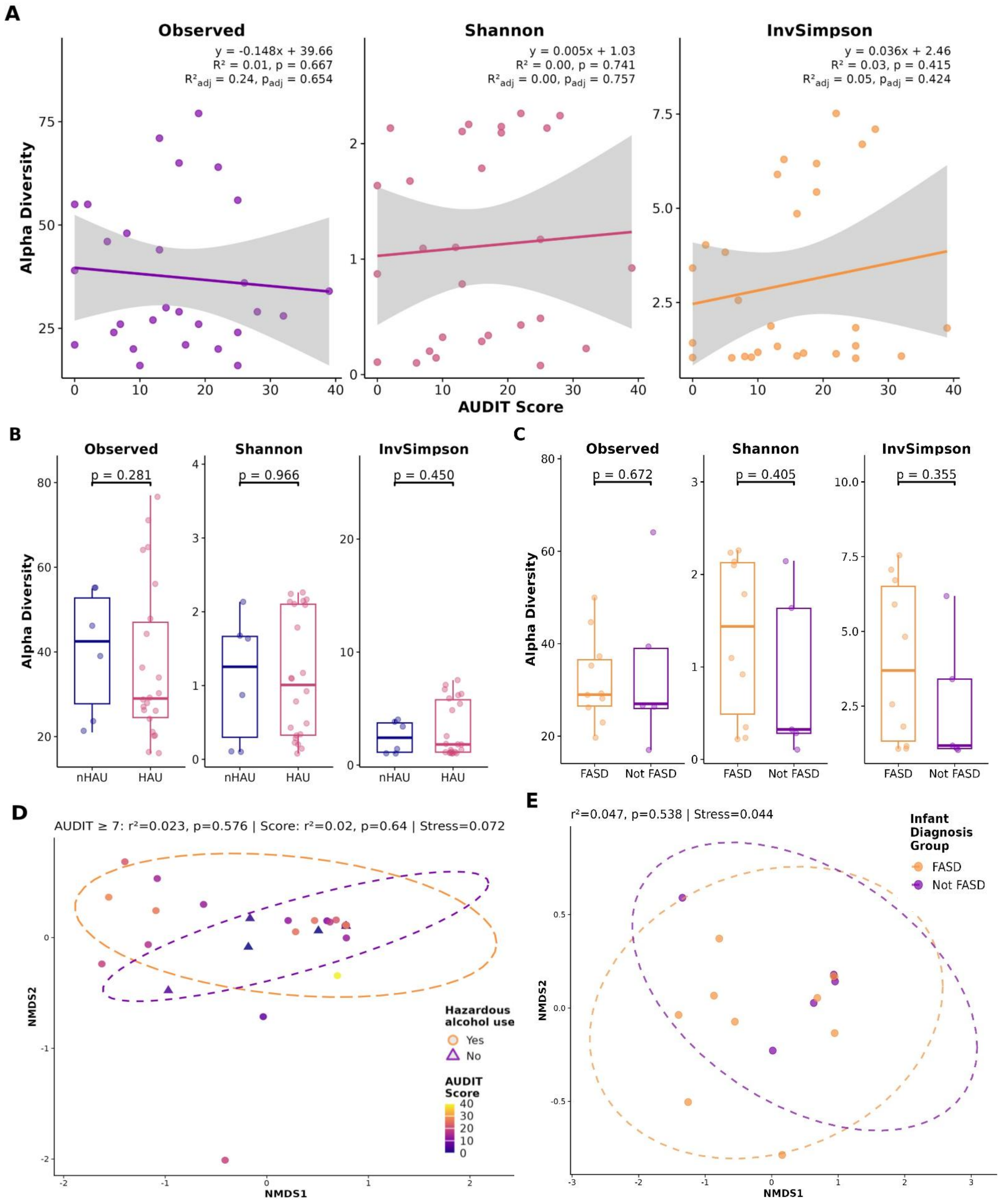


**Figure S2: Maternal vaginal microbiota diversity relative to AUDIT score and infant FASD diagnosis**. Linear regression and box-and-whisker plots of alpha-diversity indices (Observed richness, Shannon and Inverse Simpson) are shown in relation to (**A**) continuous AUDIT score (n = 28), (**B**) dichotomised AUDIT score reflecting HAU during pregnancy (AUDIT ≥ 7 : n = 22; AUDIT < 7: n = 6) and (**C**) infant FASD diagnostic group (FASD: n = 10; Not FASD: n = 5). NMDS Bray-Curtis dissimilarity ordination plots are shown for (**D**) investigated alcohol use measures, coloured by AUDIT score and stratified by alcohol use group, and (**E**) infant FASD diagnostic group. AUDIT – Alcohol Use Disorders Identification Test; FASD – fetal alcohol spectrum disorders; HAU – hazardous alcohol use; NMDS – non-metric multidimensional scaling.

#### Supporting methods: Bioinformatic analysis

Raw paired-end sequence reads were filtered using the Divisive Amplicon Denoising Algorithm 2 (*DADA2*) v.1.32.0 (Callahan et al., 2016) package using the following parameters for the gut microbiota analysis: truncLen=c(250,220), maxN=0, maxEE=c(3,3), truncQ=2, rm.phix=TRUE. The following parameters were used in the vaginal microbiota analysis: truncLen=c(240,210), maxN=0, maxEE=c(3,4), truncQ=2, rm.phix=TRUE. Filtered reads were further dereplicated, denoised, and merged using default *DADA2* parameters. Taxonomic assignment was performed using the *DADA2*-formatted SILVA SSU rRNA reference database v.138.2 (Callahan et al., 2016; Quast et al., 2013).

For the fecal microbiota analyses, a feature table comprising of 41,591 amplicon sequence variants (ASVs) derived from 207 fecal samples, with an average read length of ~ 356 bp, was constructed. For the vaginal microbiota analysis, a second independent feature table consisting of 3,846 ASVs across 28 samples, with read lengths averaging ~ 351 bp, was created. ASV tables were separately merged into a unified object with their corresponding taxonomic classifications and relevant participant metadata using *phyloseq* v.1.48.0 (McMurdie and Holmes, 2013). Sequences that remained unassigned or were not classified as “Bacteria” at kingdom-level, and those receiving “Chloroplast” or “Mitochondria” classifications at order and family level, respectively, were removed. Following filtering, 41,485 ASVs in the fecal sample dataset and 2,968 ASVs in the vaginal swab sample dataset were retained for downstream analysis. Taxa were agglomerated and investigated at the genus-level rank.

Rarefaction curves were generated using the *rarecurve* function of *vegan* (Dixon, 2003) v.2.6-10 to determine whether the minimum sequencing depth across the samples adequately captured the bacterial diversity within each dataset. The minimum read depth was found to be adequately representative of diversity, as evidenced by the plateauing of the rarefaction curves across the majority of the samples (**Figure S1**). The datasets were normalised by random sub-sampling without replacement using the *rarefy* function of the *phyloseq* (McMurdie and Holmes, 2013) package to an even depth of 18,665 and 27,331 reads for the fecal microbiota alcohol use and FASD-related analyses, and 60,667 and 81,236 reads for the vaginal microbiota alcohol use and FASD-related analyses, respectively.

Covariate adjustment sets were selected for each analysis on the basis of their independent association/s with alpha- and beta-diversity measures, statistically significant differences of variables between groups ($p \leq 0.05$) and their biological relevance. Metadata variables with too few observations for reliable statistical analysis in datasets with limited sample sizes were excluded from models. Linear regression models and permutational ANOVAs (PERMANOVAs), with 999 permutations, were used to test covariate associations with alpha-diversity metrics and beta-diversity distance matrices, and derive *p*-values and effect sizes, respectively. Covariates were retained for adjustment in downstream analyses if significantly associated with a diversity metric ($p \leq 0.05$) or if they explained ≥ 2% of variance in beta diversity ($R^2 \geq 0.02$). Covariate adjustment sets are detailed in **Table S1**.

Microbial diversity measures were generated using *vegan* (Dixon, 2003). Alpha diversity was assessed using observed richness, Shannon, and inverse Simpson diversity metrics. Differences in alpha diversity metrics between groups for the alcohol-related fecal and vaginal swab sample analysis were assessed using robust linear regression models, accounting for relevant covariate adjustment sets.

Bray-Curtis dissimilarity matrices were constructed using the metaMDS function in vegan (Dixon, 2003) and non-metric multi-dimensional scaling (NMDS) plots were generated to visualize the microbial community structure. PERMANOVAs (9,999 permutations) were performed to determine whether variables of interest were associated with differences in microbial community composition.

MaAsLin3 (v.0.99.16) (Nickols et al., 2026) was used to test associations between metadata variables and microbial community features. Analyses were run on non-rarefied data using default package parameters, with a minimum prevalence threshold of 10% and a minimum relative abundance of 0.001. In addition to the relevant covariate set for each analysis, extraction batch was modelled as a random effect and read depth was included as a covariate to account for differential sequencing depth across samples. All analyses used a two-sided test of hypothesis, a significance level of $p \leq 0.05$, and an adjusted *p*-value following Benjamini-Hochberg correction for multiple testing termed *q*. A modest significance threshold $q \leq 0.10$, chosen to enable a broader view of associations as has been performed in similar studies (White et al., 2009), was used. Genera with a nominal *p*-joint ≤ 0.05 were reported on in the absence of FDR-significant results.

*PICRUSt2* (v.2.6.1) (Douglas et al., 2020) was applied to the ASV sequences of the non-rarefied gut microbiota dataset to predict gene family abundances and the related functional pathways of the bacterial communities present. KEGG ortholog (KO) abundances were converted to KEGG pathway abundances using *ggpicrust2* (Yang et al., 2023) v.2.5.10, and MetaCyc pathway abundances were directly obtained from the *PICRUSt2* pathway inference module. Functional analysis was only performed for the fecal microbiota analysis, as functional inference tools have demonstrated poor performance when applied to vaginal samples (Carter et al., 2023). Differential abundance testing of the KEGG and MetaCyc pathways was performed using the default abundance model of *MaAsLin3*, with a minimum prevalence threshold of 10% and a minimum relative abundance of 0.001. The *p*-values were adjusted using Benjamini-Hochberg correction procedures, with $q < 0.10$ considered statistically significant and $p < 0.05$ reported as nominally significant for the purposes of this exploratory study.
